# In utero human cytomegalovirus infection expands NK cell-like FcγRIII-expressing CD8+ T cells that mediate antibody-dependent functions

**DOI:** 10.1101/2023.09.08.23295279

**Authors:** Eleanor C. Semmes, Danielle R. Nettere, Ashley N. Nelson, Jillian H. Hurst, Derek Cain, Trevor D. Burt, Joanne Kurtzberg, Keith Reeves, Carolyn B. Coyne, Genevieve Fouda, Justin Pollara, Sallie R. Permar, Kyle M. Walsh

## Abstract

Human cytomegalovirus (HCMV) profoundly modulates host T and natural killer (NK) cells across the lifespan, expanding unique effector cells bridging innate and adaptive immunity. Though HCMV is the most common congenital infection worldwide, how this ubiquitous herpesvirus impacts developing fetal T and NK cells remains unclear. Using computational flow cytometry and transcriptome profiling of cord blood from neonates with and without congenital HCMV (cCMV) infection, we identify major shifts in fetal cellular immunity marked by an expansion of Fcγ receptor III (FcγRIII)-expressing CD8+ T cells (FcRT) following HCMV exposure in utero. FcRT cells from cCMV-infected neonates express a cytotoxic NK cell-like transcriptome and mediate antigen-specific antibody-dependent functions including degranulation and IFNγ production, the hallmarks of NK cell antibody-dependent cellular cytotoxicity (ADCC). FcRT cells may represent a previously unappreciated effector population with innate-like functions that could be harnessed for maternal-infant vaccination strategies and antibody-based therapeutics in early life.

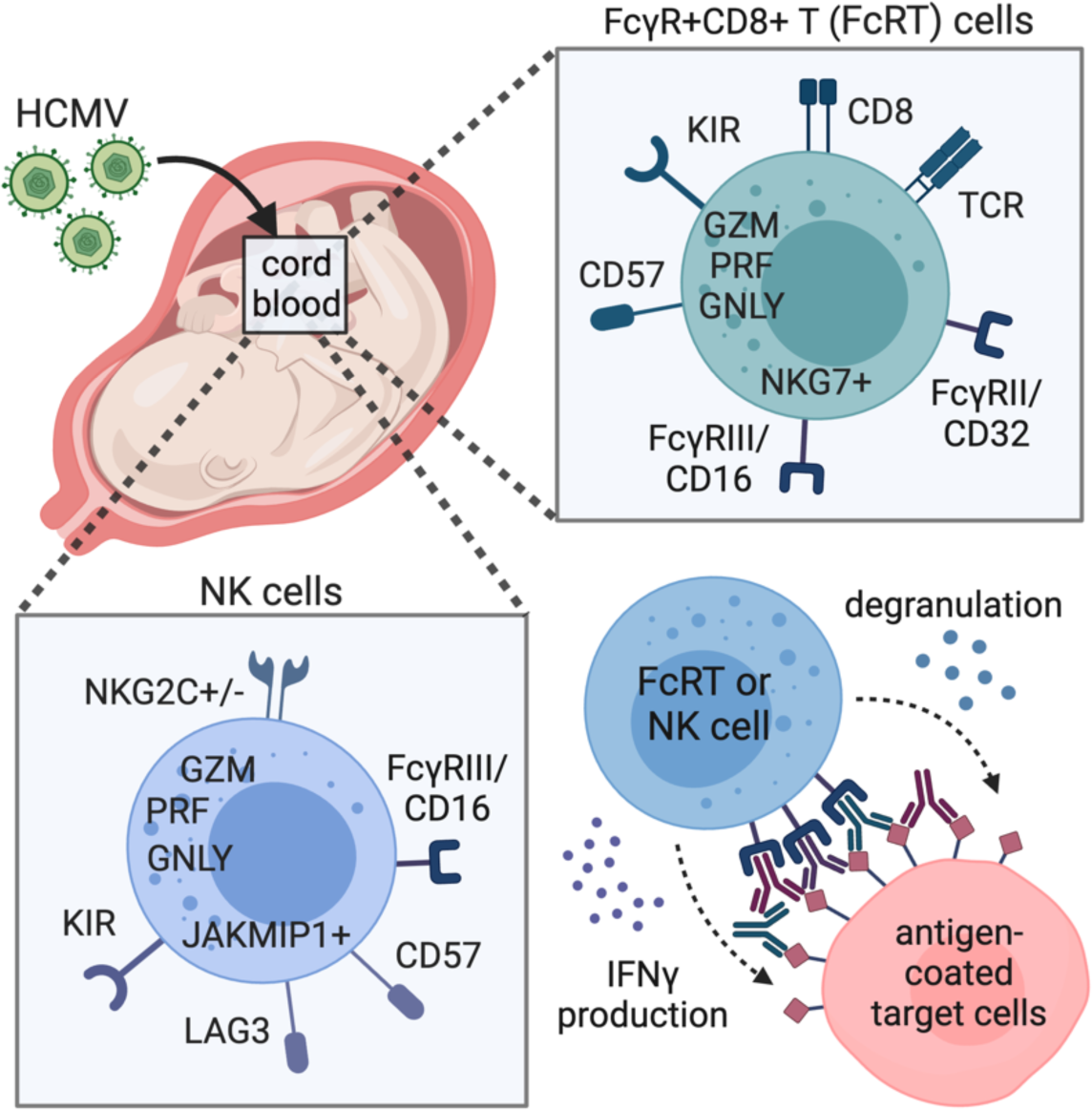

## Introduction

Human cytomegalovirus (HCMV) is a ubiquitous β-herpesvirus that has co-evolved with humans over millennia and is an important member of the human virome, a dynamic network of commensal and pathogenic viruses (Cadwell, 2015; Davis and Brodin, 2018). Globally, greater than 80% of individuals are latently infected with HCMV (Zuhair et al., 2019) and few human pathogens are known to exert such a profound imprint on host immunity across the lifespan (Brodin et al., 2015). While primary infection, latency reactivation, and reinfection are usually asymptomatic in healthy children and adults, HCMV can cause severe disease in immunocompromised populations including fetuses, transplant recipients, and persons living with HIV/AIDS. HCMV is the most common vertically transmitted infection worldwide and can cause devastating neurologic disease, yet the majority of infants born with HCMV or infected postnatally are asymptomatic (Boppana et al., 2013). Latent HCMV infection in healthy children and young adults may even enhance heterologous immune responses to infections and vaccines (Davis and Brodin, 2018; Furman et al., 2015; Semmes et al., 2020b). Thus, while HCMV remains a significant pathogen for prenatal and immunocompromised populations, emerging evidence suggests that HCMV may enhance host immunity in young, healthy individuals.

HCMV infection shapes global immune cell profiles, not just HCMV-specific cells, creating long-lasting shifts in natural killer (NK) and T cell compartments and expanding unique effector populations bridging innate and adaptive immunity (Brodin *et al*., 2015; Rolle and Brodin, 2016). “Memory-like” or “adaptive” NK cells generated by interactions between the HCMV peptide UL40 and NKG2 killer lectin-like (KLR) receptors are persistently expanded in HCMV-infected individuals and can mediate enhanced anti-viral responses upon restimulation (Guma et al., 2004; Schlums et al., 2015) . Additionally, HCMV seropositivity has been associated with a dramatic clonal expansion of HCMV-specific CD8+ T cells and with the activation and terminal differentiation of bystander non-HCMV specific CD8+ T cells (Almanzar et al., 2005; Sylwester et al., 2005). Mature CD8+ T cell subsets that express NK receptors including Fcy receptor III (FcyRIII, also known as CD16), NKG2C, and killer-like immunoglobulin receptors (KIRs) that demonstrate hybrid T-NK cell functions have also been observed in adults with chronic HCMV infection (Remmerswaal et al., 2019; Sottile et al., 2021).

Despite HCMV’s significant impact on the adult immune system, our understanding of how HCMV modulates NK and T cells in early life is limited. Fetal HCMV-specific CD8+ and CD4+ T cells and restricted γδ T cell subsets can expand following infection (Antoine et al., 2012; Huygens et al., 2014; Huygens et al., 2015; Marchant et al., 2003; Vermijlen et al., 2010), but the global impact of HCMV on developing T cells is unknown. Vaaben et al. recently reported that fetal NK cells from infants with cCMV infection highly express markers of maturation, activation, and cytotoxicity (Vaaben et al., 2022), yet the functional capacity of these NK cells and whether adaptive-like NK cell subsets are generated in utero remains unclear (Vaaben *et al*., 2022). Importantly, the fetal and neonatal immune landscape is fundamentally distinct from the adult immune system, as it is relatively biased towards immunotolerance and innate immune responses (Kollmann et al., 2017; Semmes et al., 2020a), leading us to question how HCMV exposure in utero influences developing T and NK cells.

In this study, we investigated how HCMV impacts fetal T and NK cell populations using banked cord blood from U.S. donors with and without cCMV infection. We comprehensively characterized cord blood T and NK cells using multiparameter flow cytometry, whole transcriptome profiling, and functional assays, and identified a striking expansion of a population of CD8+ T cells expressing NK cell associated markers, including FcyRIII, in cCMV-infected neonates. FcγRIII+CD8+ T cells (which we refer to as FcRT cells) expressed an NK-like transcriptional profile and mediated robust antibody-dependent functions. These findings suggest that fetal CD8+ T cells can be stimulated to differentiate into innate-like cells that mediate antibody-dependent cellular cytotoxicity (ADCC), an Fc effector function traditionally associated with NK cells. FcRT cells may have promising translational potential as a novel effector cell population linking innate and adaptive immunity that could be harnessed by vaccines and antibody-based therapeutics in early life.

## Results

### Cord blood donor immunophenotyping data highlights distinct immune landscape in cCMV-infected versus uninfected neonates

In this study, we analyzed umbilical cord blood from the U.S.-based Carolinas Cord Blood Bank (CCBB), an FDA licensed (DUCORD), public cord blood bank housed at Duke University. In the CCBB donor database, we retrospectively identified 59 cases of cCMV infection (based on screening for cord blood HCMV DNAemia) with banked biospecimens and phenotyping data available (**Supplementary Fig. 1**). Using infant sex, race/ethnicity, maternal age, and delivery year as matching variables, cCMV-infected neonates were matched to at least two control donors without cCMV infection. Demographic and clinical characteristics were similar between cCMV-infected (cCMV+; n=59) and uninfected (cCMV-; n=135) donors (**Supplementary Table 1**).

First, we analyzed cord blood donor immunophenotyping data from the CCBB donor database to identify differences between cCMV+ and cCMV-cord blood units. Total numbers and proportions of hematopoietic stem cells (CD34+), mature leukocytes (CD45+), and major immune cell lineages including 18 total immune parameters were available for analyses. There was a significant lymphocytosis in the cord blood of cCMV+ versus cCMV-neonates (*P* < 0.0001, **Fig. 1A**), characterized by a higher proportion of CD3+ T cells in cCMV+ neonates (*P* < 0.0001, **Fig. 1B**). The ratio of CD4+/CD8+ T cells was decreased in cCMV+ versus cCMV-neonates (*P* < 0.0001), driven by an expansion of CD8+ T cells (**Fig. 1C-E**). To a lesser degree, CD4-CD8-“double negative” T cells (**Fig. 1F**) and NK cells (**Fig. 1G**) were also expanded in cCMV+ neonates. Next, we used principal components analysis (PCA) to visualize the cord blood immunophenotyping data by cCMV status. Together, PC1 and PC2 accounted for ∼50% of the total variance between donors (**Fig. 1H**). Cord blood immunophenotypes from cCMV+ and cCMV-neonates clustered distinctly (**Fig. 1H**), with increased CD8+ T cells as the top parameter associated with cCMV infection (**Fig. 1I**). We also visualized the PCA by infant sex, infant race/ethnicity, and delivery mode and found no evidence that these characteristics were underlying differences in cord blood immune profiles (**Supplementary Fig. 2**). Overall, these data indicate that HCMV stimulates the expansion of fetal innate (i.e., NK cells) and adaptive (i.e., CD8+ T cells) populations.

**Figure 1.**
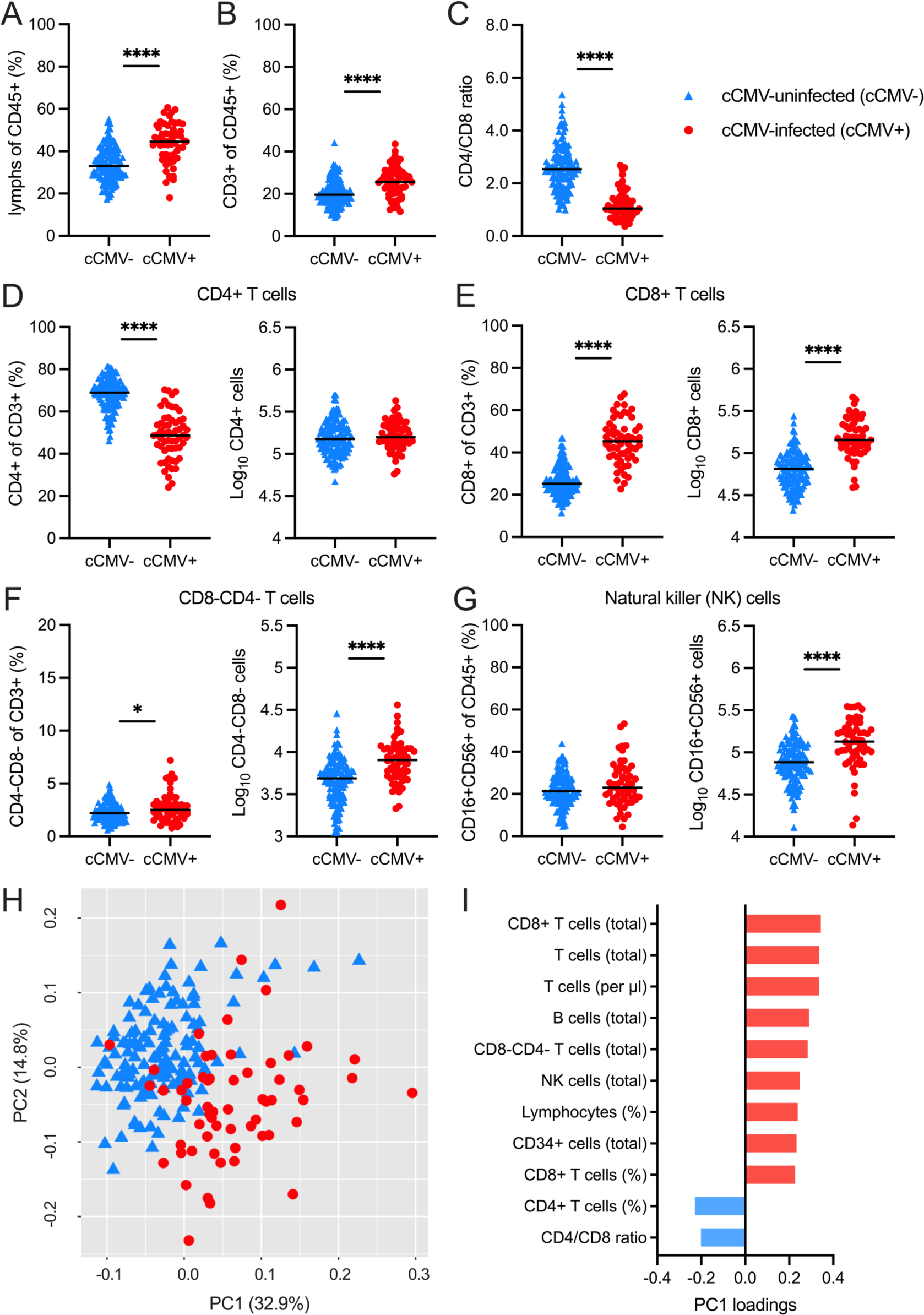
Umbilical cord blood donor cell phenotyping highlights distinct immune landscape in cCMV-infected versus uninfected neonates. Flow cytometry analysis of umbilical cord blood from n=59 cCMV-infected (cCMV+, red circles) and n=135 cCMV-uninfected (cCMV-, blue triangles) neonates was performed by the CCBB at the time of donation. (A-G) Frequencies and total immune cell counts from cord blood graft characterization. (H-I) Principal components analysis (PCA) of graft characterization data including 18 immune cell parameters from CCBB database. (H) Scatterplot of PC1 and PC2. (I) Immune cell parameter loading variables ordered by magnitude of contribution to PC1. FDR-corrected *P* values for Mann-Whitney U test. \*\*\*\**P* < 0.0001. \**P* < 0.05.

### CD56^neg^CD16/FcγRIII^+^ and NKG2C^+^ NK cells expand in cord blood from cCMV-infected versus uninfected neonates

To explore how HCMV exposure in utero influences host immune cell compartments, we performed multiparameter flow cytometry and transcriptional profiling of NK and T cells in an available subset of cord blood samples from cCMV+ (n=21) and cCMV-(n=20) neonates. Total NK cells and 3 major NK cell subsets including CD56^neg^CD16^+^, CD56^bright^CD16^+/-^, and CD56^dim^CD16^+/-^ NK cells (Ty et al., 2023; Vaaben *et al*., 2022) were compared in cCMV+ versus cCMV-neonates (**Fig. 2A**). CD56^neg^CD16^+^ NK cells were significantly expanded (**Fig. 2B**) in cCMV+ infants, and both CD56^neg^CD16^+^ and CD56^bright^CD16^+/-^ NK cells had higher expression of the activation and differentiation marker CD57 (**Fig. 2C**). NKG2C, but not NKG2A, was also more frequently expressed on NK cells from cCMV+ versus cCMV-neonates (**Fig. 2D-E**). Expression of NKG2C and CD57 are both hallmarks of the adaptive NK cell subsets expanded in adults with chronic HCMV infection (Rolle and Brodin, 2016). Next, we performed RNA sequencing of FAC-sorted total NK cells (n=13 cCMV+, n=12 cCMV-). Differential gene expression analysis identified 75 upregulated and 77 downregulated genes in NK cells from cCMV+ versus cCMV-groups (**Fig. 2F-G**) with enriched gene ontology pathways including innate immune response (*P_adj_* = 1.0×10^−4^), defense response to virus (*P_adj_* = 2.5×10^−4^), and type I interferon signaling (*P_adj_* = 6.7×10^−4^). Expression of LAG3, a checkpoint inhibitor induced by type I IFN that is highly expressed on ADCC-mediating CD56^neg^CD16^+^ NK cells (Ty *et al*., 2023), was 4-fold higher in NK cells from cCMV+ versus cCMV-neonates (*P*_FDR_ = 2.53×10^−10^). JAKMIP1, a marker of adaptive NK cells in chronic HCMV infection (Rückert et al., 2022) was also elevated 5-fold (*P*_FDR_ = 2.53×10^−3^). Together, these data demonstrate that NK cells with an anti-viral transcriptional program and adaptive phenotype expand in utero following HCMV infection.

**Figure 2.**
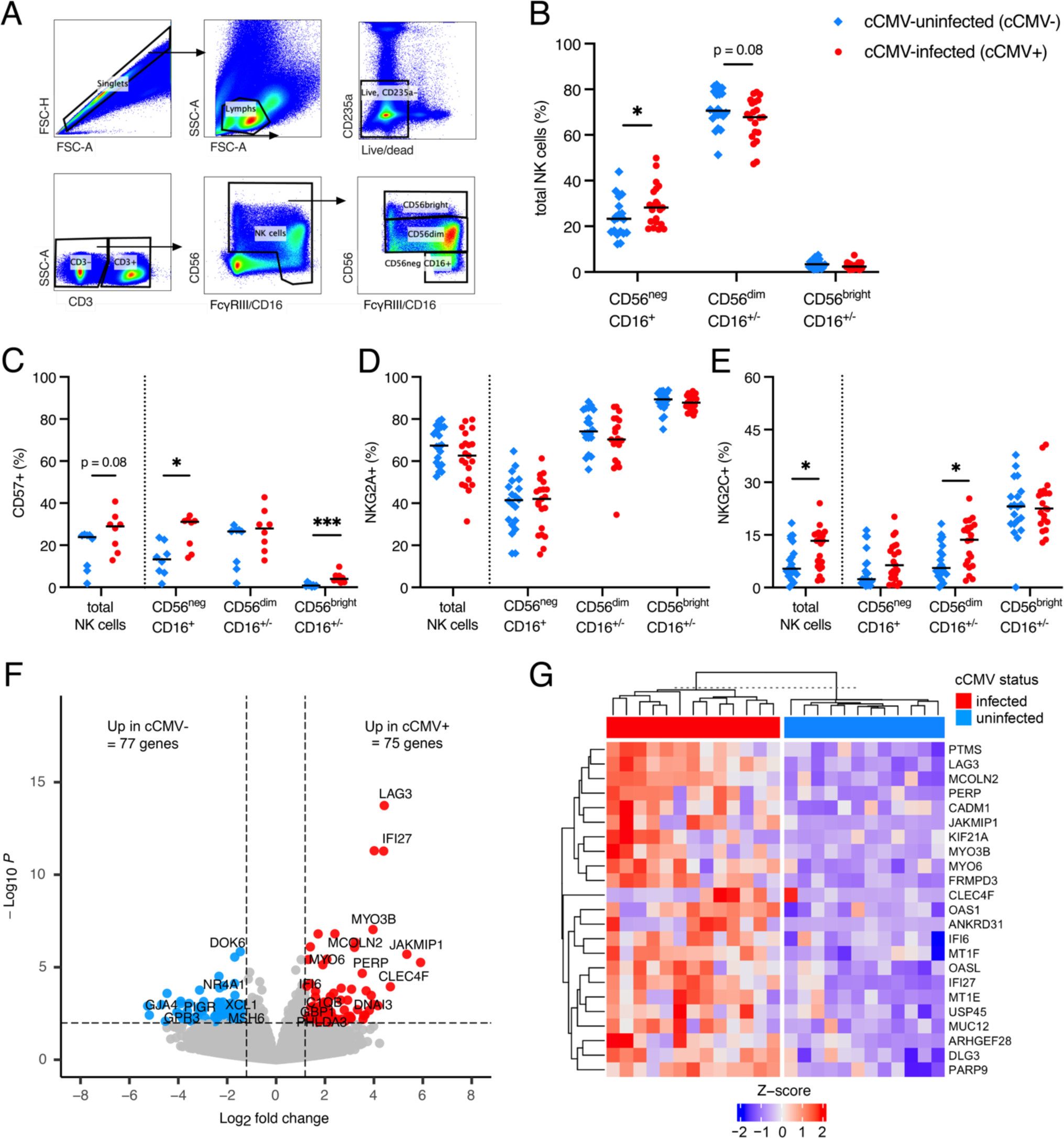
CD16/FcγRIII^+^CD56^neg^ and NKG2C^+^ NK cells expand in cord blood from cCMV-infected versus cCMV-uninfected neonates. NK cell immunophenotypes and transcriptional profiles were compared in umbilical cord blood from cCMV-infected (cCMV+, red circles) versus cCMV-uninfected (cCMV-, blue diamonds) neonates. (A) NK cell gating strategy. (B) Frequencies of NK cell subsets in cCMV+ (n=21) versus cCMV-(n=20) neonates. (C) Frequency of total NK cells and NK cell subsets expressing CD57 in cCMV+ (n=8) versus cCMV-(n=8) neonates. (D-E) Frequency of total NK cells and NK cell subsets expressing (D) NKG2A and (E) NKG2C in cCMV+ (n=21) versus cCMV-(n=20) neonates. (F) Volcano plot demonstrating differentially expressed genes in FAC-sorted total NK cells from cCMV+ (n=13) and cCMV-(n=12) neonates. Significance was set at *P* <0.01 and log2foldchange +/-1.2. Red circles indicate genes enriched in cCMV+ NK cells, blue circles indicate genes enriched in cCMV-NK cells, and grey circles indicate genes whose expression did not differ significantly between groups. (G) Heatmap of top 23 enriched genes (FDR *P* <0.1, log2foldchange >1.2) in NK cells from cCMV+ (n=13) versus cCMV-(n=12) neonates. Z-score shows gene expression based on rlog-transformed data. FDR-corrected *P* values for Mann-Whitney U test. \**P* < 0.05, \*\**P* < 0.01, \*\*\**P* < 0.001.

### CD8+ T cells upregulate cytotoxic molecules and NK cell associated genes in cord blood from cCMV-infected versus uninfected neonates

Next, we compared CD4+ and CD8+ T cells in cord blood from cCMV+ (n=21) and CMV-(n=20) neonates. There were no differences in the proportions of naïve, central memory (Tcm), effector memory (Tem), terminally differentiated effector memory T cells re-expressing CD45RA+ (Temra), or regulatory (Treg) CD4+ T cell subsets between cCMV+ and cCMV- neonates (**Supplementary Fig. 3A-B**). The frequency of CD4+ T cells expressing activation (HLA-DR, CD57), differentiation (CD57), and antigen stimulation (PD-1) markers were higher in cCMV+ infants, but these subsets were low frequency overall (**Supplementary Fig. 3C**). Differential gene expression analysis identified 180 upregulated and 368 downregulated genes in CD4+ T cells from cCMV+ (n=11) versus cCMV-(n=12) neonates (**Supplementary Fig. 3D**), yet only 25 genes remained significantly enriched after false discovery rate (FDR) correction (*P*_FDR_ < 0.01) for multiple comparisons (**Supplementary Fig. 3E**). Expression of CCL5 (*P*_FDR_ = 1.10×10^−8^), a proinflammatory chemokine that recruits T and NK cells, and natural-killer gene 7 (NKG7, *P*_FDR_ = 1.97×10^−6^), which helps traffic cytotoxic vesicles to the immunological synapse were induced 5-fold (**Supplementary Fig. 3D-E**). Transcripts for cytolytic molecules granzyme H and granzyme B were also upregulated 7-fold (*P*_FDR_ = 5.8 x 10^-6^) and 5-fold (*P*_FDR_ *=* 0.01) respectively (**Supplementary Fig. 3D-E**). Taken together, these data suggest that HCMV activates a minor subset of fetal CD4+ T cells, particularly those with cytotoxic activity or function in the recruitment of cytotoxic cells.

In contrast to the moderate changes in CD4+ T cells, we observed profound differences in the CD8+ T cell compartment. Proportions of Tcm and Tem CD8+ T cells were increased, whereas naïve CD8+ T cells were decreased in cCMV+ neonates (**Fig. 3A**). CD57 was expressed on most CD8+ T cells from cCMV+ (median = 59.4%) versus <1% of CMV-infants (**Fig. 3B**). Differential gene expression analysis from whole transcriptome sequencing identified 774 upregulated and 420 downregulated genes in CD8+ T cells from cCMV+ (n=13) versus cCMV-(n=11) groups (**Fig. 3C-D**). Multiple proinflammatory chemokines including CCL3 (*P*_FDR_ = 1.1×10^−18^), CCL4 (*P*_FDR_ = 2.1×10^−30^), and CCL5 (*P*_FDR_ = 1.7×10^−20^) were upregulated (**Fig. 3D**). Gene expression of cytolytic molecules granzyme H (*P*_FDR_ = 1.6×10^−11^), granzyme B (*P*_FDR_ = 3.8×10^−18^), perforin (*P*_FDR_ = 9.2×10^−12^), granulysin (*P*_FDR_ = 2.7×10^−14^) and NKG7 (*P*_FDR_ = 1.5×10^−24^) were increased 3-to-5-fold in cCMV+ versus cCMV-infants (**Fig. 3C-F**). Expression of transcripts encoding FcγRIIIa/CD16A (Log2FC = 5.9, *P*_FDR_ = 8.8×10^−26^), FcγRIIIb/CD16B (Log2FC = 6.9, *P*_FDR_ = 1.7×10^−13^), and killer cell lectin-like receptors (KLR) were also markedly increased in CD8+ T cells from cCMV+ neonates (**Fig 3. C-D, F**). Gene set enrichment analysis (GSEA) demonstrated that the top induced pathways in CD8+ T cells from cCMV+ versus cCMV-infants included NK cell mediated immunity (*P*_adj_ = 1.3×10^−3^), NK cell mediated cytotoxicity (*P*_adj_ = 1.3×10^−3^), and regulation of NK cell mediated immunity (*P*_adj_ = 2.9×10^−3^) (**Supplementary Table 2**). Together, these data indicate that fetal CD8+ T cells exposed to HCMV in utero have high cytotoxic potential and upregulate NK cell associated genes that may contribute to unique anti-viral functions.

**Figure 3.**
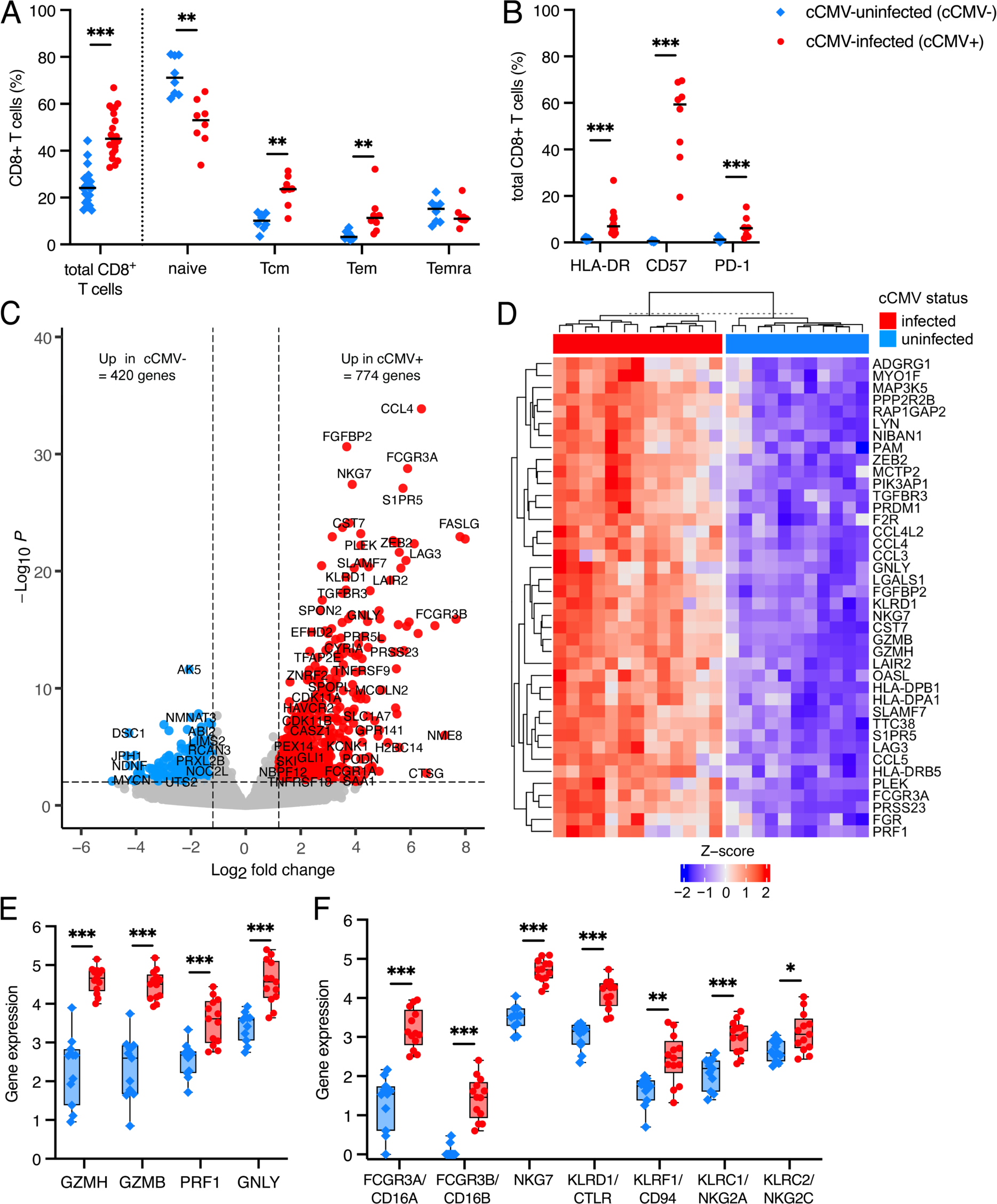
CD8+ T cells upregulate cytotoxic molecules and NK cell associated genes in cord blood from cCMV-infected versus uninfected neonates. (A-B) CD8+ T cell immunophenotypes were compared in umbilical cord blood from cCMV-infected (cCMV+, red circles) versus cCMV-uninfected (cCMV-, blue diamonds) neonates. (A) Frequency of total, naïve, central memory (Tcm), effector memory (Tem), and terminally differentiated effector memory cells re-expressing CD45RA+ (Temra) CD8+ T cells in cord blood from cCMV+ (n=21 total) versus cCMV-neonates (n=20 total). (B) Frequency of total CD8+ T cells expressing HLA-DR, CD57, and PD-1. (C) Volcano plot demonstrating differentially expressed genes in FAC-sorted total CD8+ T cells from cCMV+ (n=13) and cCMV-(n=11) neonates. Significance was set at *P* < 0.01 and log2foldchange +/-1.2. Red circles indicate genes enriched in cCMV+ CD8+ T cells, blue circles indicate genes enriched in cCMV-CD8+ T cells, and grey circles indicate genes whose expression did not differ significantly between groups. (D) Heatmap of top 40 enriched genes (FDR *P* <0.1, log2foldchange >3.0, mean count >500) in CD8+ T cells from cCMV+ (n=13) versus cCMV-(n=11) neonates. Z-score shows gene expression based on rlog-transformed data. (E-F) Selected gene expression levels in CD8+ T cells from cCMV+ (n=13) versus cCMV-(n=11) neonates including genes encoding (E) T and NK cell cytotoxic molecules and (F) NK cell associated genes. FDR-corrected *P* values for Mann-Whitney U test. \**P* < 0.05, \*\**P* < 0.01, \*\*\**P* < 0.001.

### CD8+ T cells expressing FcγRIII and NKG2A/C expand in cord blood from cCMV-infected versus uninfected neonates

To further define CD8+ T cell populations expressing NK cell markers, we analyzed our flow cytometry data using a machine learning algorithm called CITRUS (cluster identification, characterization, and regression). CITRUS uses unsupervised hierarchical clustering of multiparameter flow cytometry data instead of manual gating to identify immune cell populations that differ between groups (Bruggner et al., 2014). We first used t-SNE-CUDA (Chan et al., 2019) to visualize our 16-color single cell data (**Fig. 4A**) and select florescent channels for downstream analysis. CD3, CD4, CD8, CD127, CD25, CD19, CD56, FcγRIII/CD16, NKG2A, NKG2C, HLA-DR, and CD14 marker expression on 1,250,000 cells (50,000 cells/sample, n = 25, minimum cluster size = 1,250 cells) were used to generate the CITRUS cluster map (**Fig. 4B, Supplementary Fig. 4**). CITRUS identified 38 immune cell clusters that differed significantly (*P*_FDR_ < 0.01) between cCMV+ and cCMV-groups, including multiple clusters of activated CD8+ and CD4+ T cells that we previously identified with manual gating (**Fig. 4B**). Two clusters in the CD8+ T cell “branch”, one co-expressing NKG2A and NKG2C (**Fig. 4C**) and the other co-expressing FcγRIII and NKG2C (**Fig. 4D**), were also significantly expanded in cCMV+ versus cCMV-cord blood. These populations clustered distinctly from the NK cell “branch” (**Fig. 4B**) and had high expression of the T cell lineage marker CD3 (**Fig. 4C-D**). Using manual gating (**Fig. 4E**), we confirmed that CD8+ T cells expressing FcγRIII were significantly increased in cord blood from cCMV+ (median = 11.2%) versus cCMV-(median = 2.2%) neonates (**Fig. 4F**). The frequency of CD8+ T cells expressing NKG2A and NKG2C was also increased in cCMV+ infants and nearly absent (median <1%) in cCMV-infants (**Fig. 4F**). Together, these data demonstrate that CD8+ T cells expressing the NK cell associated receptors FcγRIII and NKG2A/C expand following HCMV exposure in utero.

**Figure 4.**
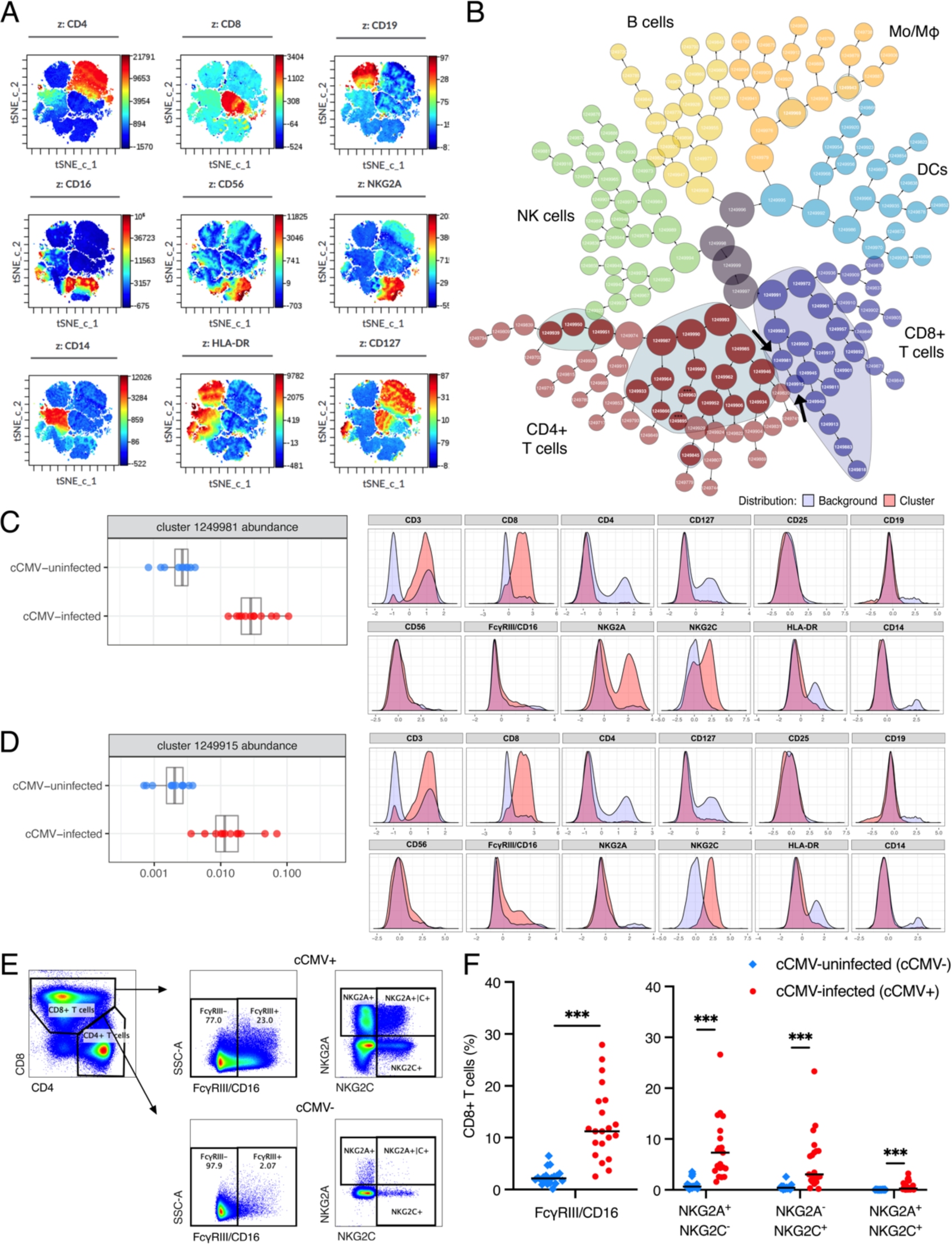
CD8+ T cells expressing NK cell receptors FcγRIII/CD16 and NKG2A/C expand in cord blood from cCMV-infected versus uninfected neonates. (A-D) Cluster identification, characterization, and regression (CITRUS) analysis of flow cytometry data was used to identify immune cell populations i.e., clusters with differing abundance in cord blood from cCMV-infected (n=13) versus cCMV-uninfected (n=12) neonates. (A) t-SNE-CUDA dimensionality reduction of flow cytometry data prior to CITRUS analysis. (B) CITRUS cluster map of 1,250,000 live, CD235a-singlet events (50,000 events/sample, minimum cluster size 1%) with ellipses indicating clusters that differed significantly (FDR *P* <0.01) between cCMV+ and cCMV-groups. Clusters colored by immune cell lineage including CD8+ T cells (purple), CD4+ T cells (red), NK cells (green), B cells (yellow), monocytes/macrophages (MO/Mϕ; orange) and dendritic cells (DCs; blue) based on marker expression in Supplementary Fig 4. (C-D) Select clusters (arrows in panel B) of CD8+ T cells expressing NK cell markers. Dot plots indicate cluster abundance in cCMV+ (red circles) versus cCMV-(blue circles) neonates. Histograms indicate fluorescent marker expression of select cluster (red) relative to background expression (blue). (E) Gating strategy to identify CD8+ T cells expressing NK cell markers. (F) Frequency of FcγRIII/CD16 and NKG2A/C expression on total CD8+ T cells from cCMV+ (red circles, n=21) versus cCMV-(blue diamonds, n=20) neonates. FDR-corrected *P* values for Mann-Whitney U test. \*\*\**P* < 0.001.

### FcγRIII+CD8+ T (FcRT) cells in cord blood from cCMV-infected neonates are a heterogenous population expressing activation and terminal differentiation markers

CD8+ T cells expressing NK cell markers have been described as a differentiated cell subset with NK-like functions in adults with chronic viral infections including HCMV, EBV, HIV and HCV (Björkström et al., 2008; Clémenceau et al., 2008; Mazzarino et al., 2005; Naluyima et al., 2019; Pietra et al., 2003). To further characterize these cells in cord blood, we designed a 14-color panel with select T and NK cell markers based on this prior literature. CITRUS analysis of CD3, CD4, CD8, CD56, FcγRIII (CD16), γδ TCR, CCR7, CD45RA, PD-1, CD57, NKG2A, and NKG2C marker expression on 1,200,000 cells (75,000 cells/sample, n=16 samples, minimum cluster size = 1,200 cells) generated a cluster map with distinct “branches” of T and NK cell clusters and identified 28 clusters that differed significantly (*P*_FDR_ < 0.01) between cCMV+ (n=8) and cCMV-(n=8) groups (**Fig. 5A, Supplementary Fig. 5**). Multiple CD8+ T cell clusters with increased FcγRIII expression were enriched in cCMV+ cord blood (**Fig. 5A, Supplementary Fig. 5**). Most FcγRIII+CD8+ T cell clusters resembled canonical differentiated CD8+ T cells, whereas two clusters expressed the γδ TCR and had variable expression of activation and differentiation markers (**Fig. 5B**). We next confirmed these differences by manual gating on FcγRIII+ and FcγRIII-CD8+ T cells. While the majority of FcγRIII+CD8+ T cells did not express the γδ TCR, there was a higher frequency of γδ TCR expression in the FcyRIII+ (median = 27%) versus FcγRIII-CD8+ T cell subsets (**Fig. 5C**). FcγRIII+CD8+ T cells were more likely to be Temra than FcγRIII-CD8+ T cells (**Fig. 5D**). The majority of FcγRIII+CD8+ T cells expressed CD57 (median = 77%) whereas PD-1 and NKG2A/C expression was more variable (**Fig. 5D-E**). Together, these data suggest that FcγRIII+CD8+ T cells, which we refer to as FcRT cells, are a heterogenous population of activated and terminally differentiated T cells that expand in utero following HCMV infection.

**Figure 5.**
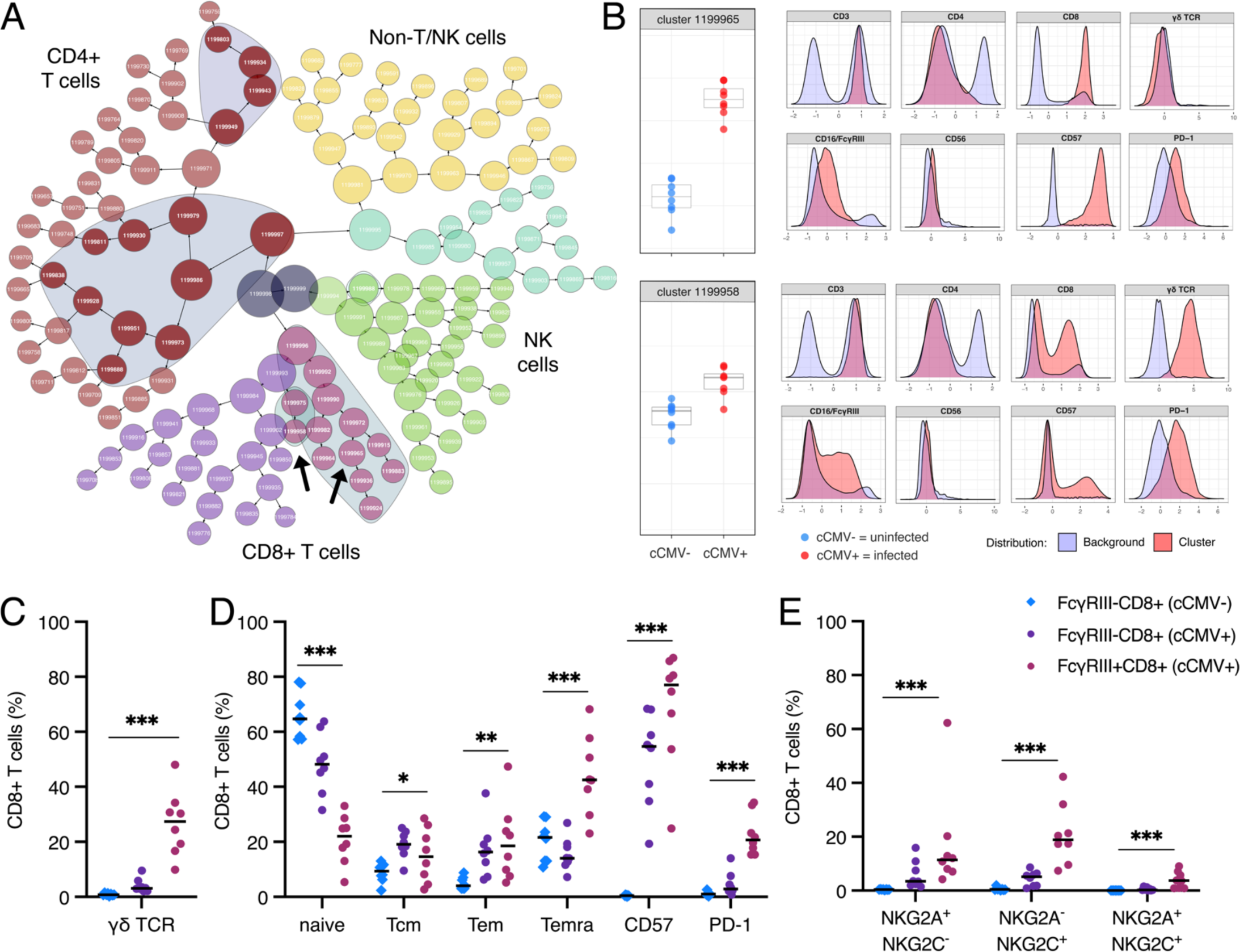
FcγRIII+ CD8+ T (FcRT) cells in cord blood from cCMV-infected neonates are a heterogenous population expressing activation and terminal differentiation markers. (A-B) Cluster identification, characterization, and regression (CITRUS) analysis of flow cytometry data was used to identify immune cell populations i.e., clusters with differing abundance in cord blood from cCMV-infected (n=8) versus cCMV-uninfected (n=8) neonates. (A) CITRUS cluster map of 1,125,000 live, CD235a-singlet events (75,000 events/sample, minimum cluster size 1%) with ellipses indicating clusters that differed significantly (FDR *P* <0.01) between cCMV-infected and uninfected groups. Clusters colored by immune cell lineage including CD8+ T cells (purple = FcγRIII-, plum = FcγRIII+), CD4+ T cells (red), NK cells (green), and non-T/NK cells (yellow, aqua) based on marker expression in Supplementary Fig 5. (B) Select clusters (indicated by arrows in panel A) of CD8+ T cells expressing NK cell associated markers. Dot plots indicate cluster abundance in cord blood from cCMV+ (red circles, n=13) versus cCMV-(blue circles, n=12) neonates. Histograms indicate fluorescent marker expression of select cluster (red) relative to background expression (blue). (C-E) Immunophenotypes of FcγRIII+ (plum) and FcγRIII-(cCMV+: purple; cCMV-: blue) CD8+ T cells from cCMV+ (circles, n=8) and cCMV-(diamonds, n=7) neonates. (C) Frequency of γδ T cell receptor expression. (D) Frequency of naïve, central memory (Tcm), effector memory (Tem), and terminally differentiated effector memory cells re-expressing CD45RA+ (Temra) phenotype based on CD45RA and CCR7. Frequency of subsets expressing CD57 or PD-1. (E) Frequency of NKG2A and NKG2C expression. *P* values for Kruskal-Wallis test \**P* < 0.05 \*\**P* < 0.01 \*\*\**P* < 0.001.

### FcγRIII+CD8+ T (FcRT) cells in cord blood from cCMV-infected neonates upregulate a NK cell-like antibody effector transcriptional program

Next, we compared the whole transcriptome of FcRT and FcγRIII-CD8+ T cells from cCMV+ and cCMV-neonates. Cytolytic molecules, chemotaxic chemokines, and sphingosine-1-phosphate receptor 5 (S1PR5), which regulates T cell infiltration and migration, were upregulated in FcRT and FcγRIII-CD8+ T cells from cCMV+ infants (**Fig. 6A-B, Supplementary Fig. 6A-B**). FcγRIII-CD8+ T cells from cCMV-infants had a distinct transcriptional profile compared to FcRT cells with increased expression of IL7R and CCR7, genes associated with naïve T cells (**Fig. 6C-D**). FcγRIII-CD8+ T cells from cCMV+ infants had an intermediate transcriptional profile that clustered between FcRT cells and FcγRIII-CD8+ T cells from cCMV-infants (**Fig. 6C-E**). KIR and KLR gene expression was highly upregulated in FcRT cells (**Fig. 6E, Supplementary Fig. 6C-D**) as were additional “NK cell identity” genes including CD244, NCR1, NCAM1, TYROBP, and ITGAX that were preselected based on prior publications (Naluyima *et al*., 2019; Sottile *et al*., 2021) (**Fig. 6E**). PCA of T and NK cells together further demonstrated that FcRT cells acquire an NK-like transcriptional profile (**Fig. 7A**) with expression of cytotoxic molecules and FcγRs approaching or exceeding NK cells (**Fig. 7B-C**). In contrast, FcγRIII-CD8+ T cells from cCMV+ infants expressed a mixed transcriptional profile with variable induction of NK cell associated markers and FcγRs (**Fig. 6E, Fig. 7A-C**). FCGR3A, which encodes FcγRIIIa/CD16A, was the most highly expressed FcγR in FcRT cells; however, gene expression of FCGR3B (FcγRIIIb/CD16B), FCGR2B (FcγRIIb/CD32B), and FCGR2C (FcγRIIIc/CD32C), which can mediate ADCC or other Fc effector functions, was also increased (**Fig. 7C**). Together, these data demonstrate that FcRT cells in cCMV+ neonates acquire an NK cell-like transcriptional profile with the potential capacity to execute antibody-mediated cytotoxic effector functions.

**Figure 6.**
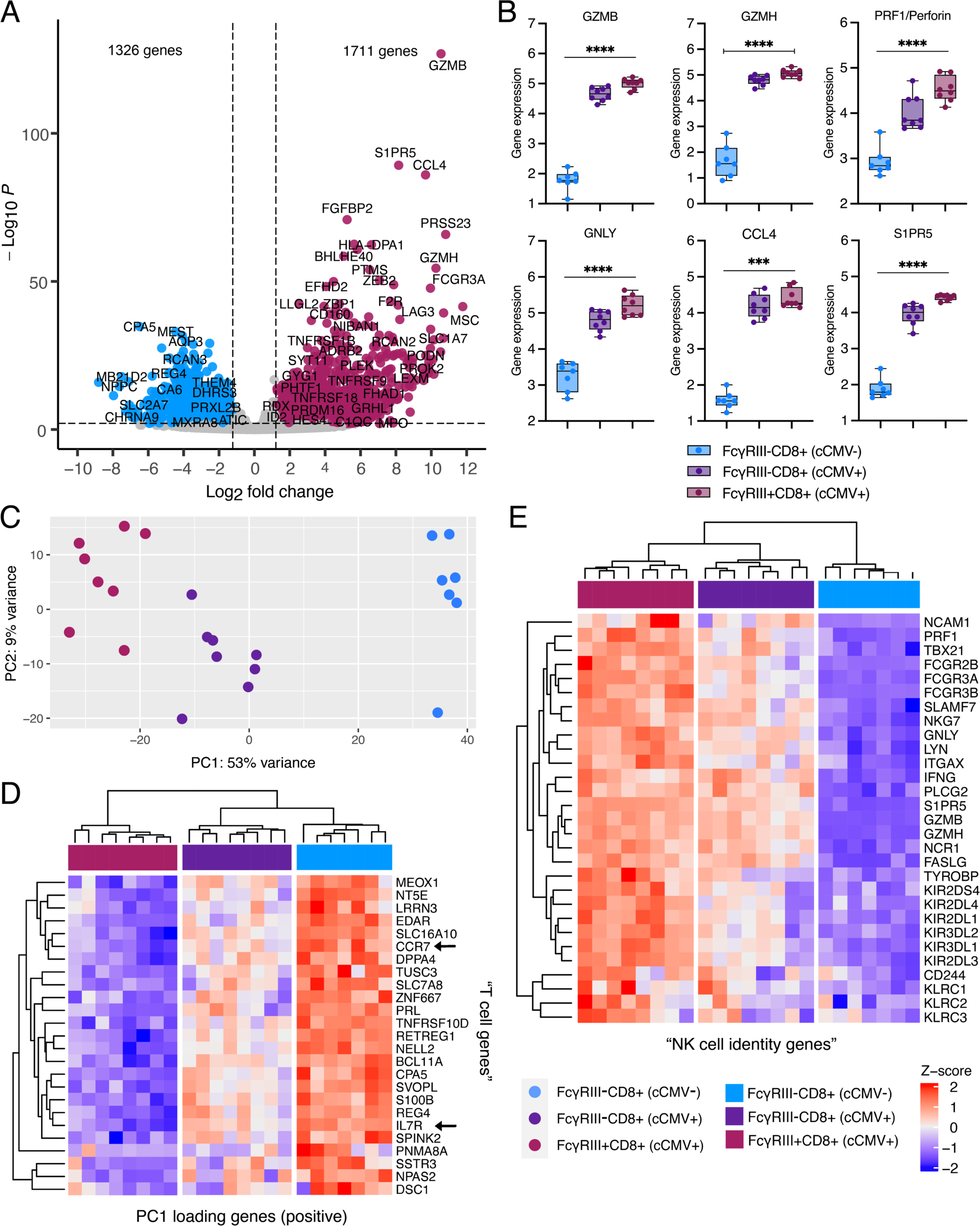
FcγRIII+CD8+ T cells (FcRT) in cord blood from cCMV-infected neonates upregulate a NK cell-like antibody effector transcriptional program. (A-E) Transcriptome analysis of FAC-sorted FcγRIII+ and FcγRIII-CD8+ T cells from cCMV-infected (n=8) and cCMV-uninfected (n=7) neonates. (A) Volcano plot demonstrating differentially expressed genes in FcγRIII+ versus FcγRIII-CD8+ T cells. Significance was set at *P* <0.01 and log2foldchange +/-1.2. Plum circles indicate genes enriched in FcγRIII+ CD8+ T cells (cCMV+ only), blue circles indicate genes enriched in FcγRIII-CD8+ T cells (cCMV-only), and grey circles indicate genes whose expression did not differ significantly between groups. (B) Selected gene expression levels in FcγRIII+ and FcγRIII-CD8+ T cells. (C) Principal components analysis (PCA) of top 500 differentially expressed genes in FcγRIII+ versus FcγRIII-CD8+ T cells. (D-E) Heatmaps of differentially expressed genes in FcγRIII+ and FcγRIII-CD8+ T cells. Z-score shows gene expression based on rlog-transformed data. (D) Heatmap of top 25 PC1 loading genes. (E) Heatmap of NK cell identity genes. *P* values for Kruskal-Wallis test \*\*\**P* < 0.001 \*\*\*\**P* < 0.001.

**Figure 7.**
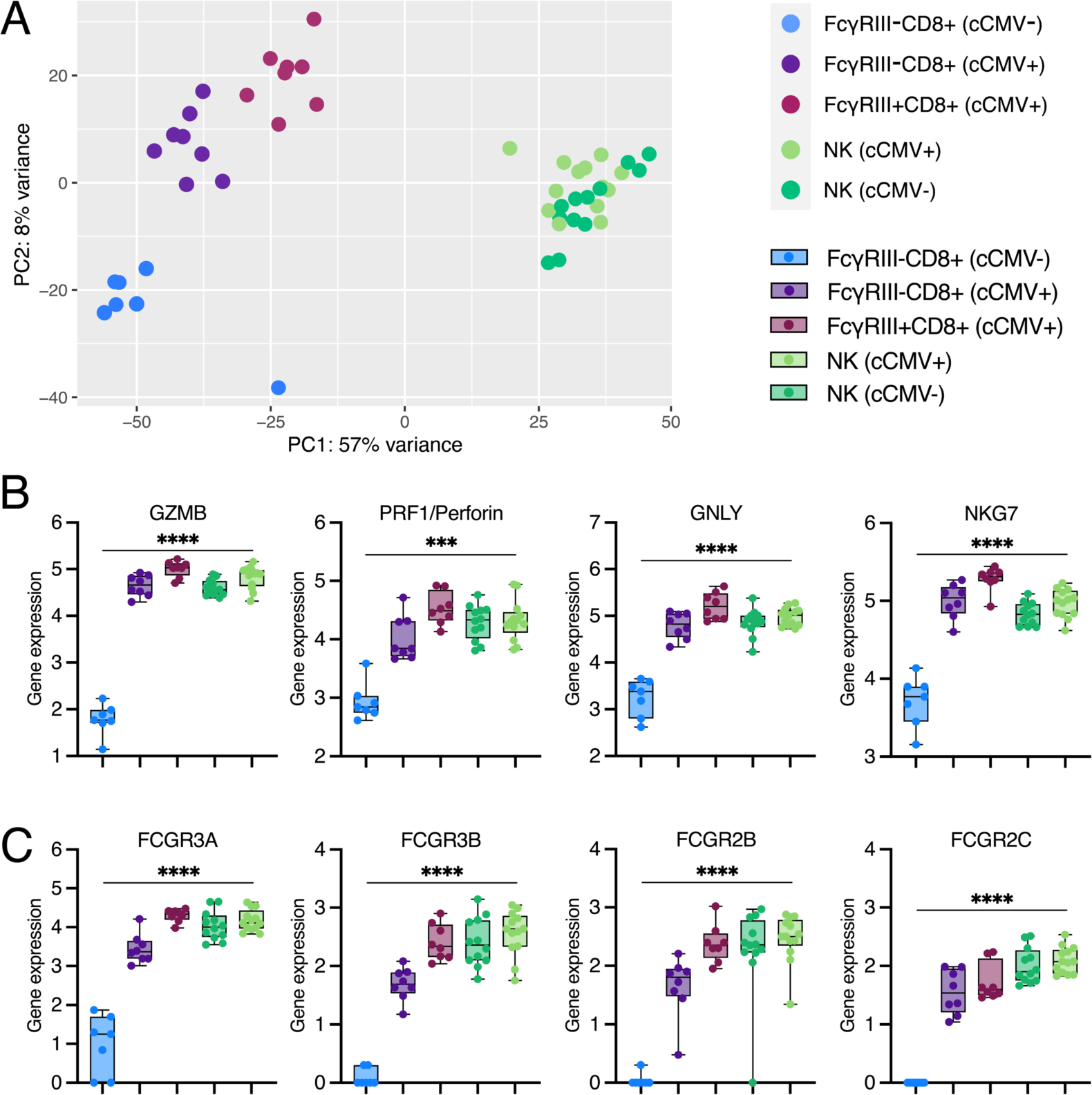
FcγRIII+ CD8+ T (FcRT) cells in cord blood from cCMV-infected neonates acquire an NK-like transcriptional profile. (A-C) Transcriptome analysis of FAC-sorted FcγRIII-CD8+ T cells, FcγRIII+ CD8+ T cells, and NK cells from cCMV-infected and uninfected neonates. (A) Principal components analysis (PCA) of top 500 differentially expressed genes in FAC-sorted CD8+ T and NK cells. (B-D) Gene expression levels of (B) cytotoxic molecules and (C) FcγR genes. *P* values for Kruskal-Wallis test \*\*\**P* < 0.01 \*\*\*\**P* < 0.001.

### Cord blood NK and FcγRIII+CD8+ T (FcRT) cells produce IFNγ and degranulate against antibody-opsonized target cells

To assess whether FcRT cells can mediate Fc-dependent effector functions, we used cord blood NK and CD8+ T cells to measure antibody-dependent degranulation and IFNγ production, which are key markers of ADCC activity (**Fig. 8A**). Using intracellular staining, we confirmed our transcriptional data showing higher protein expression of perforin and granzyme B in FcRT cells (**Fig. 8B**). The frequency of FcRT cells containing perforin and granzyme B was high (median = 85.9%) and even exceeded autologous NK cells (median = 73%) from cCMV+ neonates (**Fig. 8B**). To test FcγR function, we employed a previously-established assay for assessing NK cell ADCC activity (Chung et al., 2009). We used an HIV model system, as all donors previously screened HIV negative, ensuring responses to the challenge antigen were antibody-mediated and not influenced by memory T cell responses. First, we measured NK cell degranulation and IFNγ production in response to antibody stimulation, which was comparable in cord blood from cCMV+ and cCMV-neonates (**Fig. 8C-D**). Both markers of ADCC activity were enhanced when NK cells were preincubated with IL-15 (**Fig. 8C-D**), consistent with prior literature indicating that IL-15 augments neonatal NK cell cytotoxicity (Pollara et al., 2020). Next, we assessed the capacity of cord blood CD8+ T cells to mediate Fc-dependent antibody effector functions. FcyRIII-CD8+ T cells from cCMV+ and cCMV-infants did not degranulate or produce effector cytokines against HIV antigen coated cells when co-incubated with non-specific or HIV-specific antibodies (**Fig. 8E-H**). In contrast, FcRT cells from cCMV+ neonates were able to degranulate, as measured by CD107a expression, and produce IFNγ in an antigen-specific antibody-dependent manner (**Fig. 8E-H**). Moreover, FcRT cells demonstrated increased degranulation and IFNγ production if preincubated with IL-15 (**Fig. 8E-H**), similar to autologous NK cells (**Fig. 8C-D**). Altogether, these data demonstrate that cord blood FcRT cells as well as NK cells can mediate robust ADCC-associated antibody-dependent functions that are enhanced by effector cytokines.

**Figure 8.**
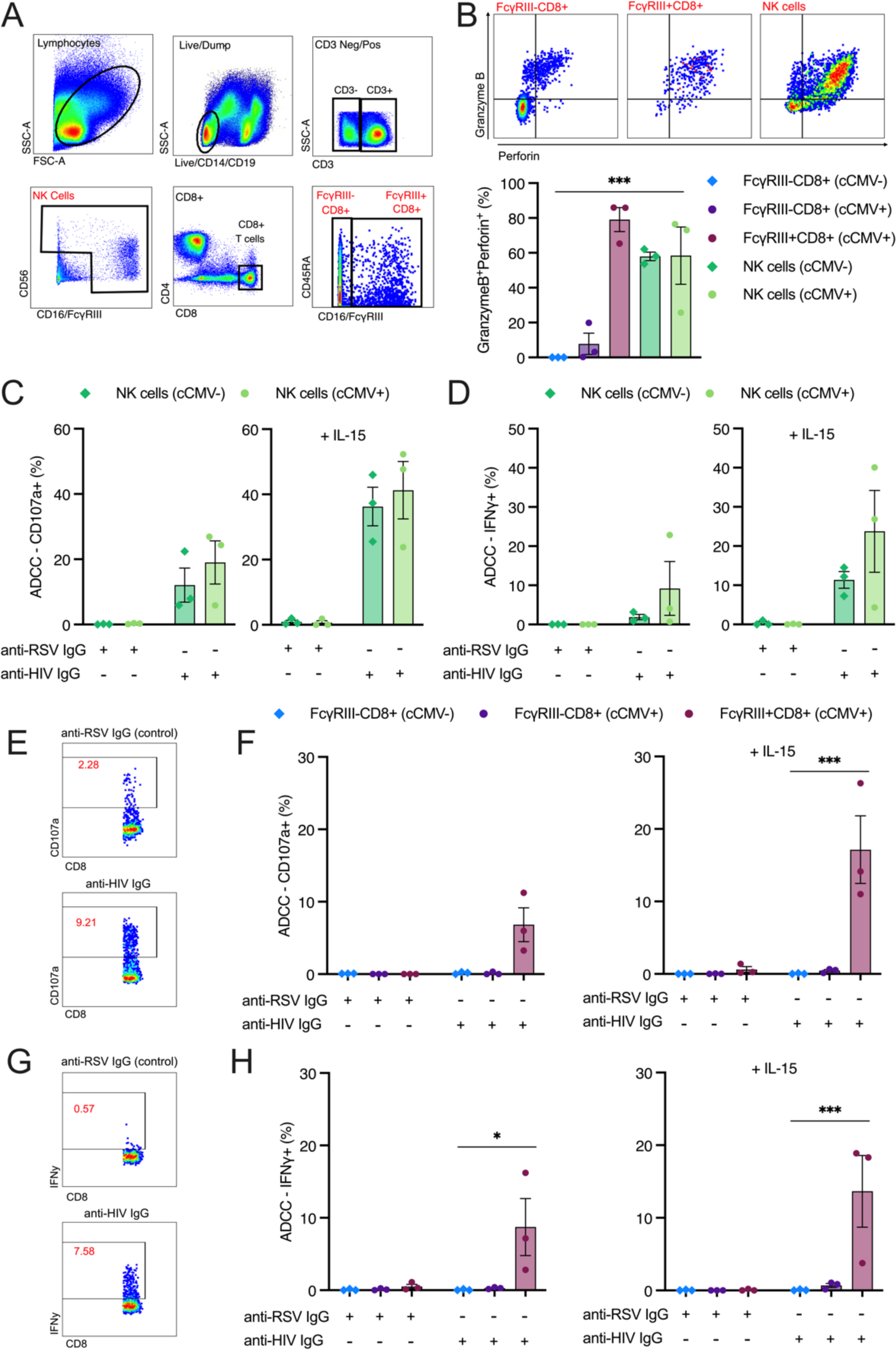
Cord blood NK and FcγRIII+CD8+ T (FcRT) cells produce IFNγ and degranulate against antibody-opsonized target cells. (A-H) Degranulation (CD107a positivity) and IFNγ production following antibody stimulation with anti-RSV IgG (non-specific antibody) or anti-HIV IgG (target cell specific antibody) were measured as markers of antibody-dependent cellular cytotoxic (ADCC) responses in cord blood NK and CD8+ T cells from cCMV-infected (n=3 circles) and uninfected (n=3 diamonds) infants. (A) Gating strategy to identify NK cells, FcγRIII+CD8+ T cells, and FcγRIII-CD8+ T cells. (B) Gating strategy to quantify granzyme B and perforin expression. Percentage of cell population co-expressing granzyme B and perforin at baseline (based on intracellular cytokine staining). (C-D) NK cell degranulation and IFNγ production following antibody stimulation with and without IL-15 pretreatment in cCMV+ (light green circles) and cCMV-(dark green diamonds) infants. (E-F) CD8+ T cell degranulation (CD107a) following antibody stimulation with and without IL-15 pretreatment. (G-H) CD8+ T cell IFNγ production following antibody stimulation with and without IL-15 pretreatment. *P* values for Kruskal-Wallis test \**P* < 0.05 \*\*\**P* < 0.001.

## Discussion

Congenital infections like HCMV pose a unique challenge to the developing immune system, which must balance the competing demands of anti-pathogen defense versus immunotolerance to maternal alloantigens, commensal microbiota, and environmental antigens (Kollmann *et al*., 2017). As such, fetal and neonatal immune cells favor innate over adaptive responses (Galindo-Albarrán et al., 2016; Semmes *et al*., 2020a) and antigen-specific responses by newly generated T and B cells remain limited. While HCMV-specific T cells have been observed in cord blood from cCMV+ infants, these adaptive T cell responses against HCMV are constrained by functional exhaustion acquired in utero (Antoine *et al*., 2012; Huygens *et al*., 2014; Huygens *et al*., 2015). Here, we demonstrate that cord blood CD8+ T cells acquire an NK-like phenotype and mediate innate effector functions through FcγRIII following HCMV exposure in utero. These results identify an alternative pathway by which the developing immune system can overcome the limitations to adaptive immunity in early life by engaging CD8+ T cells in Fc-mediated immunity. We show that FcγRIII+CD8+ T cells (which we refer to as FcRT cells) and NK cells in cord blood from cCMV-infected neonates respond robustly to antibody stimulation via degranulation and IFNγ production, indicating that both FcRT and NK cells are poised to mediate ADCC. That FcRT cells highly expressed granzyme, perforin, and granulysin and NKG-7, which facilitates cytotoxic granule exocytosis and enhances NK and CD8+ T cell cytotoxicity (Malarkannan, 2020; Ng et al., 2020), underscores the cytotoxic potential of these cells. Altogether, our work suggests that FcγRIII-expressing T and NK cells can bridge innate and adaptive immunity through Fc-mediated antibody functions and may contribute to host defense during early infant immune development.

NK-like CD8+ T cells have been described in adults with chronic viral infections including HIV (Naluyima *et al*., 2019; Phaahla et al., 2019), HCV (Björkström *et al*., 2008), EBV (Clémenceau *et al*., 2008), and HCMV (Mazzarino *et al*., 2005; Pietra *et al*., 2003; Sottile *et al*., 2021). These NK-like CD8+ T cells do not fit the characteristics of iNKT or canonical γδ T cells, but rather represent separate heterogenous subpopulations of innate-like T cells (Koh et al., 2023). Other groups have described FcyRIII-expressing CD8+ T cells in adults that can mediate ADCC (Björkström *et al*., 2008; Clémenceau *et al*., 2008; Naluyima *et al*., 2019), which can be further enhanced by IL-15 (Choi et al., 2023), Our study demonstrates that FcRT cells with a similar transcriptional profile, cytokine responsiveness, and functionality can be induced in an immature, developing immune system. Given the limited gestational window in which these infections can occur, our identification of these NK-like CD8+ T cells in cord blood challenges the assumption that these cells develop over months to years following chronic antigenic stimulation (Koh *et al*., 2023). Our findings indicate that this may be a more conserved and fundamental pathway for T cells to take given the ability of developing fetal T cells to acquire these transcriptional changes and functions in early life. That the fetal immune system can rapidly develop NK-like CD8+ T cells in response to an infectious stimulus suggests that neither chronic infection nor a fully developed adult immune system is required to generate these populations. Our work reveals that FcRT cells may be a previously unappreciated effector cell population present in early life that could contribute to host defense by acquiring antibody effector functions.

Our study also identified minor, but notable, changes in the NK cell compartment following cCMV infection. Some cord blood samples from cCMV-infected neonates had an expansion of NKG2C+ NK cells, which is a marker for the adaptive NK cells associated with chronic HCMV infection in adults and children (Guma *et al*., 2004; Guma et al., 2006; Monsivais-Urenda et al., 2010; Noyola et al., 2012). Because this process is driven by epigenetic reprogramming following repeated antigenic stimulation (Lee et al., 2015; Schlums *et al*., 2015), this may be a slower occurring adaptation, partially explaining why it was not present in all cCMV+ neonates. Rather, a more consistent difference we identified between cord blood from cCMV+ and cCMV-infants was the expansion of “atypical” or “adaptive-like” CD56neg NK cells. Expansion of CD56neg NK cells, which are believed to function through FcyRIII-mediated ADCC rather than direct cytotoxicity (Forconi et al., 2018; Forconi et al., 2020; Ty *et al*., 2023), has been observed in HCMV, EBV and malaria infection in early life (Forconi *et al*., 2018; Ty *et al*., 2023; Vaaben *et al*., 2022). CD56neg NK cells highly express CD57 and upregulate LAG3, mirroring the transcriptional changes we observed in cCMV infection, but their functionality remains unclear. Our work suggests that NK cells expanded following HCMV exposure in utero have the capacity to execute ADCC but whether CD56neg NK cells aid in anti-viral control of HCMV via antibody-dependent mechanisms should be explored in future studies (Semmes and Permar, 2022).

Our data provide important insights into the influence of cCMV infection on host cellular immunity and also highlight an opportunity to protect neonates more broadly using antibody-based interventions against infected, malignant, or autoimmune cells. Maternal IgG antibodies are actively transferred across the placenta via FcγRs to protect infants from infectious diseases (Fouda et al., 2018; Jennewein et al., 2017; Jennewein et al., 2019; Martinez et al., 2019). We speculate that FcRT cells may represent an additional effector cell population that can expand the cellular compartment to leverage maternal IgG in early life. Vaaben et al. recently proposed that FcγRIII-activating maternal IgG may synergize with neonatal NK cells to protect against HCMV via antibody-dependent mechanisms (Semmes and Permar, 2022; Vaaben *et al*., 2022), a hypothesis that our data further supports. We recently reported that higher levels of FcγRIII and ADCC-activating IgG in maternal and cord blood sera were associated with protection against fetal HCMV transmission in this same cohort (Semmes et al., 2023; Semmes et al., 2022). Thomas et al. also found that higher ADCC-mediating antibody responses and viral susceptibility to ADCC were associated with decreased risk of HIV-1 transmission in utero (Thomas et al., 2022). Together, these findings suggest that Fc-mediated responses linking maternal and fetal immunity may contribute to protection against congenital infections. We propose that both neonatal NK and FcRT cells could be targeted with therapeutic monoclonal antibodies or vaccines designed to engage both the maternal and infant immune systems.

There are several limitations to our study that could be expanded upon in future work. Our retrospective cohort limited us from collecting additional clinical data and longitudinal biospecimens. As such, we could not investigate how these changes in the cellular compartment relate to clinical outcomes or whether the gestational timing of transmission influenced fetal FcRT or NK cell responses. Moreover, we could not determine how long these changes in the cellular compartment persist and whether they maintain functionality over time. Finally, banked cord blood sample volumes were limited, so we could only perform functional studies on a subset of infants. Nevertheless, the data presented convincingly demonstrate that cCMV infection expands a CD8+ T cell subset expressing a NK cell-like phenotype, transcriptional profile, and functionality. Future studies should investigate how these immunological changes relate to anti-viral control and clinical outcomes of cCMV infection and further characterize the origin, persistence, and functions of FcRT cells in early life.

In conclusion, we have demonstrated that cCMV infection profoundly modulates developing T and NK cell compartments, marked by the expansion of FcyRIII-expressing CD8+ T cells that have high cytotoxic potential and respond robustly to antibody stimulation. The role of FcRT and NK cells in fetal defense against congenital infections must be explored further, but both cell populations represent promising translational targets to overcome the challenges to generating adaptive immune responses in early life. Altogether, our work suggests that both FcRT and NK cells can mediate antibody effector functions through FcγR engagement and could be harnessed by maternal-infant vaccination strategies or antibody-based therapeutics to protect the infant against infectious diseases.

## Methods

### Human umbilical cord blood samples

Our study included cases of congenital cytomegalovirus infection (cCMV) and controls without cCMV infection that were recruited from 2008-2017 as donors to the Carolinas Cord Blood Bank (CCBB). Approval was obtained from Duke’s Institutional Review Board (Pro00089256) to use de-identified clinical data and biospecimens provided by the CCBB. No patients were prospectively recruited for this study and all cord blood was acquired retrospectively from the CCBB biorepository from donors who provided written consent for biospecimens to be used for research. Cases and controls were identified from over 29,000 CCBB donor records (see **Supplementary Fig. 1** for an overview of sample selection). Maternal donors underwent infectious diseases screening for HCMV, hepatitis B virus, syphilis, hepatitis C virus, HIV-1/2, HTLV I and II, Chagas Disease, and West Nile virus. Only donors with healthy, uncomplicated pregnancies that gave birth at term were included and infants were screened for signs of (a) neonatal sepsis, (b) congenital infection (petechial rash, thrombocytopenia, hepatosplenomegaly), and (c) congenital abnormalities. Cord blood plasma was screened by the CCBB for HCMV viremia with a Real-Time PCR COBAS AmpliPrep/TaqMan nucleic acid test (Roche Diagnostics). Cases of cCMV infection were defined as donors with cord blood that screened positive for HCMV DNAemia per PCR. Cases with cCMV infection (cCMV+, n=59) were matched to at least 2 uninfected controls (cCMV-, n=135) that did not have detectable HCMV DNAemia in the cord blood at birth. Matching variables included infant sex, race/ethnicity, maternal age (+/-3 years), and delivery year (+/-3 years), as reported in **Supplementary Table 1**.

### CCBB graft characterization

Flow cytometry graft characterization was performed at the time of donation on fresh umbilical cord blood mononuclear cells (CBMCs) by the Duke Stem Cell Transplant Laboratory of Duke University Hospital, a CAP and FACT accredited, CLIA certified clinical laboratory which provides contract services to the CCBB. Graft characterization data was then obtained retrospectively from the CCBB donor database. PCA plots of graft characterization data were rendered using ggplot2 (v3.4.0) in R.

### NK and T cell phenotyping and sorting

Flow cytometry was performed in the Duke Human Vaccine Institute (DHVI) Research Flow Cytometry Shared Resource Facility (Durham, NC). For phenotyping, cryopreserved umbilical cord blood was thawed briefly at 37°C and resuspended in R10 media (RPM1 1640 with glutamine [Gibco] plus 10% heat-inactivated fetal bovine serum [FBS]) with Benzonase (Millipore; 2ul/mL). Re-suspended cord blood was then pelleted at 1500 rpm for 5 minutes. Following pelleting, fetal red blood cells were lysed with ∼3 mL of RBC lysis buffer for 5 mins then washed with 1X PBS and pelleted at 1500 rpm for 5 mins. CBMCs were then re-suspended and enumerated on a Muse Cell Analyzer before being pelleted at 1500 rpm for 5 mins and re-suspended at 2.0×10^7^ cells/mL in 1% PBS/BSA. For phenotyping, 5-10 million cells (depending on viable cell count after cryopreservation) were stained with an optimized monoclonal antibody cocktail of fluorescently conjugated antibodies against surface markers for 30 mins at 4°C. Antibodies in the general lineage panel included: CD14 pacific blue (M5E2, Biolegend), CD16 BV570 (3G8, Biolegend), CD25 BV605 (BC96, Biolegend), CD56 BV650 (HCD56, Biolegend), NKG2C BV711 (134591), CD45 BV786 (HI30, BD Biosciences), CD34 FITC (561, Biolegend), CD19 BB700 (HIB19, BD Biosciences), NKG2A PE (S19004C, Biolegend), CD235a PE-Cy-5 (HIR2, Biolegend), CD3 PE-Texas Red (7D6, ThermoFisher), CD127 PE-Cy7 (A019D5, Biolegend), CD8 APC (RPA-T8, BD Biosciences), HLA-DR AF700 (L243, Biolegend), and CD4 APC-H7 (SK3, BD Biosciences). Antibodies in the T and NK cell panel included: CD3 BV421 (UCHT1, BD Biosciences), CD8 BV570 (RPA-T8, Biolegend), CCR7 BV605 (G043H7, Biolegend), CD56 BV650 (HCD56, Biolegend), PD-1 BV785 (EH12.2H7, Biolegend), TCRγ/δ FITC (11F2, BD Biosciences), CD45RA PerCP-Cy5.5 (HI100, Biolegend), NKG2C PE (S19005E, Biolegend), CD57 PE-CF594 (NK-1, BD Biosciences), CD235a PE-Cy5 (HIR2, Biolegend), CD16 PE-Cy7 (3G8, BD Biosciences), and NKG2A PE-Cy5 (HIR2, Biolegend) and CD4 AF700 (L200, BD Biosciences). Cells were then washed with PBS and pelleted at 1500 rpm for 5 mins and resuspended in live/dead Aqua (ThermoFisher) or near IR (Invitrogen) stain and incubated at room temperature for 20 mins. Fluorescence minus one (FMO) control tubes were included for CD34, CD16, CD56, CD127, HLA-DR, TCR γ/δ, CCR7, CD45RA, NKG2A, NKG2C, CD57, and PD-1 for downstream manual gating. Single color AbC or ArC beads (Invitrogen) for each antibody and live/dead stain were used as compensation controls. Flow cytometry data was acquired on a FACSAria (BD Biosciences) instrument using FACSDiva (v8.0) and analyzed in FlowJo (v10.8.1).

### CITRUS analysis

tSNE-CUDA dimensionality reduction and CITRUS (cluster identification, characterization, and regression) analyses (Bruggner *et al*., 2014) were completed in Cytobank, a cloud-based bioinformatics platform for analyzing high dimensional cytometry data (Beckman Coulter; www.cytobank.org). All samples were pre-gated on live, CD235 negative cells before FCS files before downstream analyses. For the tSNE-CUDA analysis, 400,000 live, CD235-events were sampled per sample FCS file and perplexity was set to 40. For the general lineage panel CITRUS analysis, 50,000 live, CD235-events were sampled per sample FCS file (n=25 CBMCs, total cell events = 1,250,000) and the minimum cluster size was set to 1% of total events. CD3, CD4, CD8, CD127, CD25, CD19, CD56, CD16, NKG2A, NKG2C, HLA-DR, and CD14 marker expression was normalized across all samples then used as channels for clustering. For the T and NK cell panel CITRUS analysis, 75,000 live, CD235-events were sampled per sample FCS file (n=16 CBMCs, total cell events = 1,200,000) and the minimum cluster size was set to 1% of total events. CD3, CD4, CD8, CD56, CD16, γδ TCR, CCR7, CD45RA, PD-1, CD57, NKG2A, and NKG2C marker expression was normalized across all samples then used as channels for clustering. SAM, a correlative association model, was used to identify cell clusters that differed in abundance (significance cut-off FDR P < 0.01) between cCMV+ and cCMV-groups.

### RNA-seq sample preparation and analysis

T and NK cell subsets were FAC-sorted directly into RLT lysis buffer (Qiagen), and total RNA was extracted using the RNeasy Micro Kit (Qiagen Cat. No. 74004). Total CD4+ T, CD8+ T, and NK cells were sorted from 25 unique cord blood samples (n=13 cCMV+, n=12 cCMV-); however, several samples failed RNA quality control and were excluded from downstream transcriptional analyses. CD16-CD8+ and CD16+CD8+ T cell subsets were sorted from a total of 16 unique cord blood samples (n=8 cCMV+, n=8 cCMV-), and one sample from the cCMV-group failed QC and was excluded from downstream transcriptional analyses. RNA quality was evaluated by RIN number (minimum cut-off > 8.5) prior to library preparation by the Duke Human Vaccine Institute (DHVI) Sequencing Core Facility. Briefly, full length cDNAs were generated using up to 10ng of total RNA through the SMART-Seq v4 Ultra Low Input Kit for Sequencing (Takara Cat. No. 634891). Total 200pg cDNAs were used to generate the dual index Illumina libraries using Nextera XT DNA Library Prep Kit (Illumina Cat No. FC-131-1096). Sequencing was performed on an Illumina NextSeq 500 sequencer to generate 2 ’ 76 paired-end reads using TG NextSeq 500/550 High Output kit v2.5 (150 cycles) following the manufacturer’s protocol (Illumina, Cat. No. 20024912). The quality of cDNAs and Illumina libraries were assessed on a TapeStation 2200 with the high sensitivity D5000 ScreenTape (Agilent Cat, No. 5067-5592), and their quantity were determined by Qubit 3.0 fluorometer (Thermo Fisher). Gene reads were aligned to the human reference genome GRCh38 using Qiagen CLC genomics (v20). DEseq2 (v1.38.3) was used to normalize count data and perform differential gene expression analysis. Genes were considered differentially expressed based on a 1.2 log2fold change in gene expression and FDR *P* < 0.1. PCA was performed on rlog-transformed data using the plotPCA function in DESeq2. Volcano plots were generated using the EnhancedVolcano (v1.16.0) package in R. Heatmaps and hierarchical clustering was performed on rlog-normalized DEseq2data using the ComplexHeatmap (2.14.0) package in R. Gene set enrichment analysis of differentially expressed gene ontology (GO) pathways (min 5, max 2000 genes) was performed using iDEP v0.96 (Ge et al., 2018) with a significance cut-off of FDR P < 0.2.

### Functional immunological assays

Antibody-dependent degranulation of NK and CD8+ T cells was performed as follows. Cord blood was thawed at 37°C, diluted 1:4 in RPMI supplemented with 10% FBS, penicillin, streptomycin, L-glutamine, and gentamicin (complete media) and processed using Ficoll separation. Following separation, the cells were counted and rested overnight at a concentration 2-3 million cells per mL in either complete media or complete media supplemented with 1ng/mL of IL-15. After resting overnight, the cells were counted and resuspended at 5 million/mL. CBMCs in bulk are referred to as “Effector” cells and CEM.NKRs coated with 5 ug/mL BAL gp120 are referred to as “Target” cells. CBMCs were either plated alone (Effector only), with a 10:1 ratio with targets (Effector+Target), and with 1ug/mL of Synagis (Effector+Targets+Synagis) or 1ug/mL a mixture of 4 optimized HIV antibodies (Effector+Target+mAb Mix). The HIV antibody cocktail comprised of 250 ng/mL each of 7B2_AAA, 2G12_AAA, A32_AAA, and CH44_AAA which contain the AAA optimization for Fc mediated activity. All four conditions were plated for 6 hours in the presence of 1ug/mL brefeldin (BD GolgiPlug), 1ug/mL Monensin A (BD GolgiStop, and anti-CD107a antibody (Biolegend H4A3). After 6 hours, the cells were washed with DPBS and stained with Live/Dead viability stain (ThermoFischer), then washed and stained for the following surface markers: CD14 V500 (M5E2, BD biosciences), CD19 V500 (HIB19, BD biosciences), CD57 BV711 (QA17A04, Bioelegend), CD4 BV785 (OKT4, Biolegend), CD45RA FITC (5H9, BD biosciences), CD16 PE (3g8, Biolegend). CCR7 Pe-Cy5 (G043H7, Biolegend), CD8 PerCP Cy5-5 (SK1, Biolegend), CD56 PeCy7 (NCAM16.2, BD biosciences), CD3 APC-Cy7 (SK7, BD biosciences). Next, cells were fixed with CytoFix/CytoPerm (BD Biosciences) then stained for the following intracellular markers: Perforin PacBlue (dg9, Biolegend), IFNγ APC (4S.B3, Biolegend), Granzyme B AF700 (QA16A02, Biolegend) in the presence of Perm/Wash Buffer (BD Biosciences). Samples were acquired on a LSRFortessa II (BD biosciences) using FACSDiva v8.0 software. Frequency of NK and CD8+ T cells expressing granzyme B and perforin were measured in the effector only condition. Percentage change in IFNγ and CD107a were calculated by subtracting the frequencies in the Effector+Target conditions from the antibody conditions. Data was analyzed using FlowJo Version 10.8.

### Statistical Analysis

All statistical analyses were completed in R (v4.1.1) or GraphPad Prism (v9.1). Frequencies of immune cell populations or normalized gene expression data were compared using Mann-Whitney U/Wilcoxon rank-sum tests for pair-wise comparisons and Kruskal-Wallis one-way ANOVA for comparisons across multiple groups. Statistical significance was defined a prior as *P* < 0.05 with a two-tailed test and FDR correction for multiple comparisons. Specific details on each statistical analysis performed and the exact n are available in the respective figure legends. Additional details on the statistical significance thresholds for the CITRUS and RNA-seq analyses are described in the methods section above. Inclusion and exclusion criteria for the study are described above and outlined in **Supplementary Fig. 1** and randomization for case-control matching was achieved using a random number generator.

## Author contributions

Conceptualization, ECS, DRN, GF, JP, SRP, KMW; Formal Analysis, ECS, DRN; Methodology, ECS, TDB, CBC, KR, JP, SRP, KMW; Data curation, ECS; Funding acquisition, ECS, CBC, JHH, SRP, KMW; Investigation, ECS, DRN, AN; Project administration, JHH; Resources, DC, KR, JK, CBC; Software, ECS, CBC; Supervision, JP, SRP, KMW; Visualization, ECS, DRN; Writing – original draft, ECS, DRN; Writing – review & editing, ECS, DRN, AN, JHH, DC, TDB, JK, KR, CBC, GF, JP, SRP, KMW.

## Data Availability

All data produced in the present study are available upon reasonable request to the authors.

## Acknowledgements

Thank you to the CCBB donors and staff including Jose Hernandez, Ann Kaestner, and Korrynn Vincent who were instrumental in acquiring the biospecimens and donor clinical information for this study. We also would like to thank Aria Arus-Altuz and Evan Trudeau at the Duke Human Vaccine Institute Flow Cytometry Facility (Durham, NC) for their assistance in panel design and FAC-sorting, and Yue Chen and Bhavna Hora at the Duke Human Vaccine Institute Sequencing Core for their help in with RNA sequencing. This project was supported by NIH NCI 1R21CA242439-01 “Immune Correlates and Mechanisms of Perinatal Cytomegalovirus Infection and Later Life ALL Development” (KMW, SRP), NIH NIAID 1R01AI173333 “Identifying and modeling immune correlates of protection against congenital CMV transmission after primary maternal infection” (SRP), NIH NIAID R01AI145828 “Innate immune signaling in placental antiviral defenses” (CBC), the Triangle Center for Evolutionary Medicine (TriCEM) graduate student research award “Human cytomegalovirus and host B cell evolution across the lifespan” to ECS and the Translating Duke Health Children’s Health and Discovery Initiative. The funders had no role in study design, data collection and analysis, decision to publish, or preparation of the manuscript.

