## Supplementary Figures and Tables for "In utero human cytomegalovirus infection expands NK cell-like FcγRIII-expressing CD8+ T cells that mediate antibody-dependent functions"

**Supplementary Figure 1.** Identification of cases and controls from the Carolinas Cord Blood Bank (CCBB) biorepository.

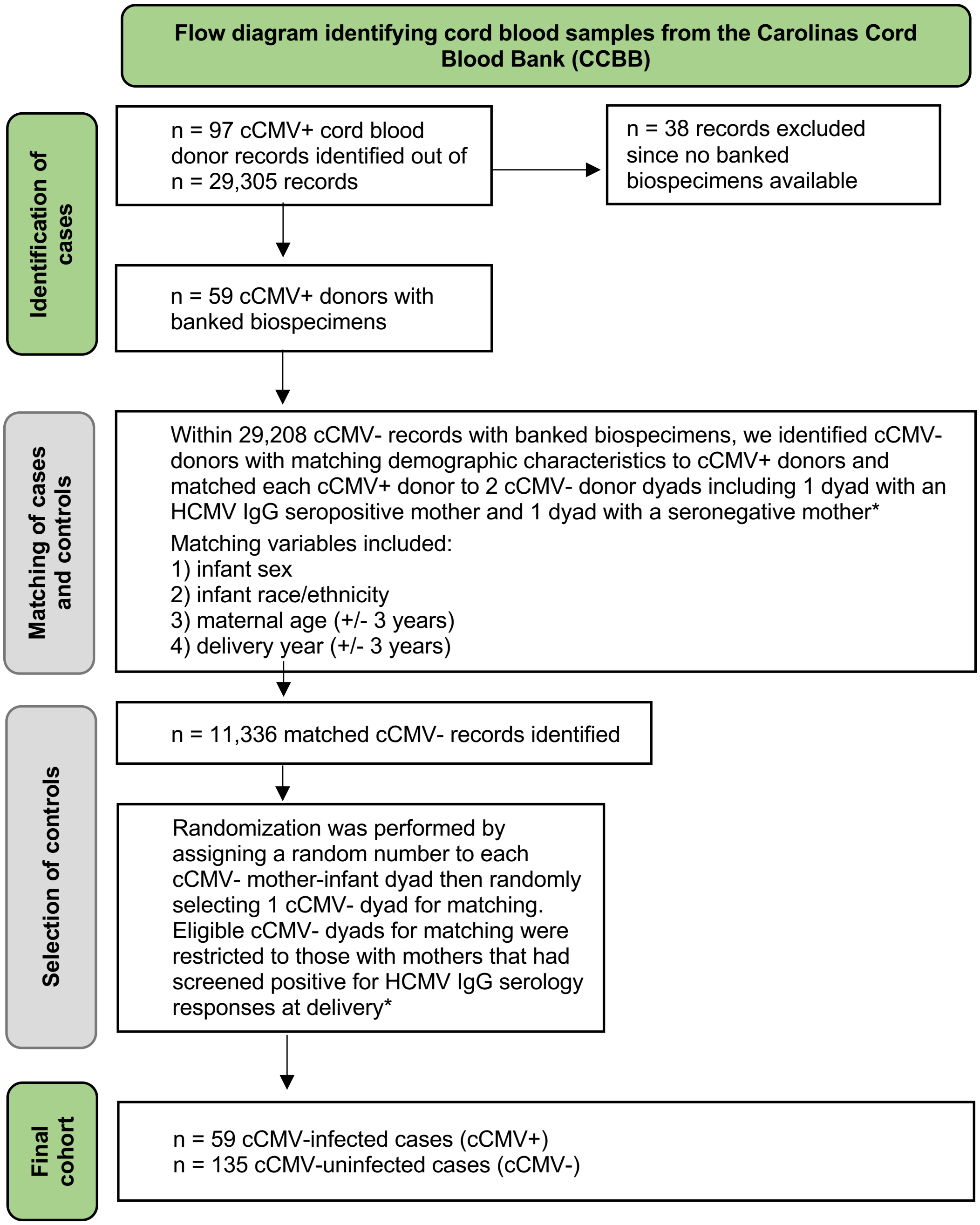


cCMV = congenital cytomegalovirus (CMV) infection
cCMV+ = positive CMV PCR cord blood screening at birth
cCMV- = negative CMV PCR cord blood screening at birth

*Initial HCMV serology screening performed at time of donation by the American Red Cross in Charlotte, N.C.

**Supplementary Table 1. Maternal and neonatal cord blood bank donor clinical characteristics[a]**

| | cCMV+ (n = 59) | cCMV- (n = 135) |
|---|---|---|
| **Infant sex, n (%)** | | |
| **Female** | 24 (40.7) | 52 (38.1) |
| **Male** | 35 (59.3) | 83 (61.9) |
| **Infant race/ethnicity, n (%)** | | |
| **White** | 32 (54.2) | 75 (55.6) |
| **Black** | 14 (23.7) | 28 (20.7) |
| **Hispanic** | 7 (11.9) | 16 (11.9) |
| **Multiple** | 1 (1.7) | 0 (0.0) |
| **Other** | 5 (8.5) | 16 (11.9) |
| **Maternal age (years), median [IQR]** | 27 [22-31] | 30 [27-34] |
| **Gestational age (weeks), median [IQR]** | 39 [38-40] | 39 [39-40] |
| **Delivery year, median [IQR]** | 2011 [2009-2015] | 2014 [2010-2017] |
| **Delivery type, n (%)** | | |
| **Vaginal** | 23 (39.0]) | 75 (55.6) |
| **Cesarean section** | 36 (61.0) | 60 (44.4) |
| **Maternal HCMV IgG seropositivity, n (%)** | | |
| **Seropositive** | 59 (100) | 0 (0.0) |
| **Seronegative** | 0 (0.0) | 135 (100) |
| **Cord blood HCMV viral load, median [range] [b]** | 727 [137-18,100] | ND |

cCMV = cCMV infection; ND = not detected

cCMV-infected (cCMV+) indicates mother-infant pair with cCMV infection

cCMV-uninfected (cCMV-) indicates mother-infant pair without cCMV infection

[a] cCMV+ and cCMV- mother-infant pairs were matched on maternal age (+/- 3 years), infant race, sex, and delivery year (+/- 3 years)

[b] Cord blood HCMV viral copies listed in IU/mL, lower limit of detection = 137 copies/mL (n = 41)

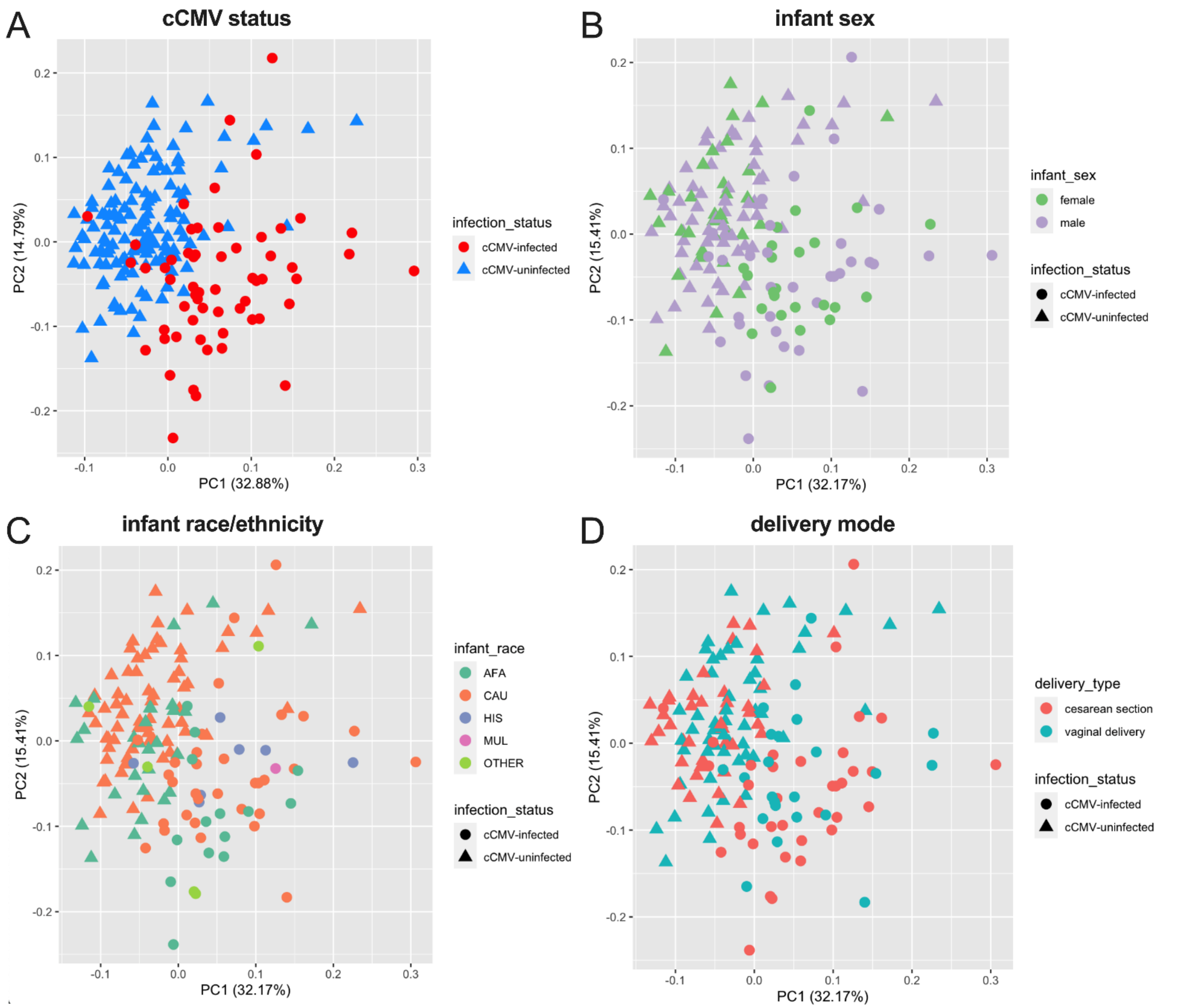


**Supplementary Figure 2. Principal components analysis (PCA) of umbilical cord blood from cCMV-infected compared to uninfected neonates stratified by clinical characteristics.** PCA of 18 umbilical cord blood immune variables from CCBB cord blood graft characterization prior to banking. (A) PCA colored by cCMV infection status. (B) PCA colored by infant sex. (C) PCA colored by infant race/ethnicity. (D) PCA colored by delivery mode. n = 59 cCMV-infected (cCMV+, circles), n = 135 cCMV-uninfected (cCMV-, triangles) neonates.

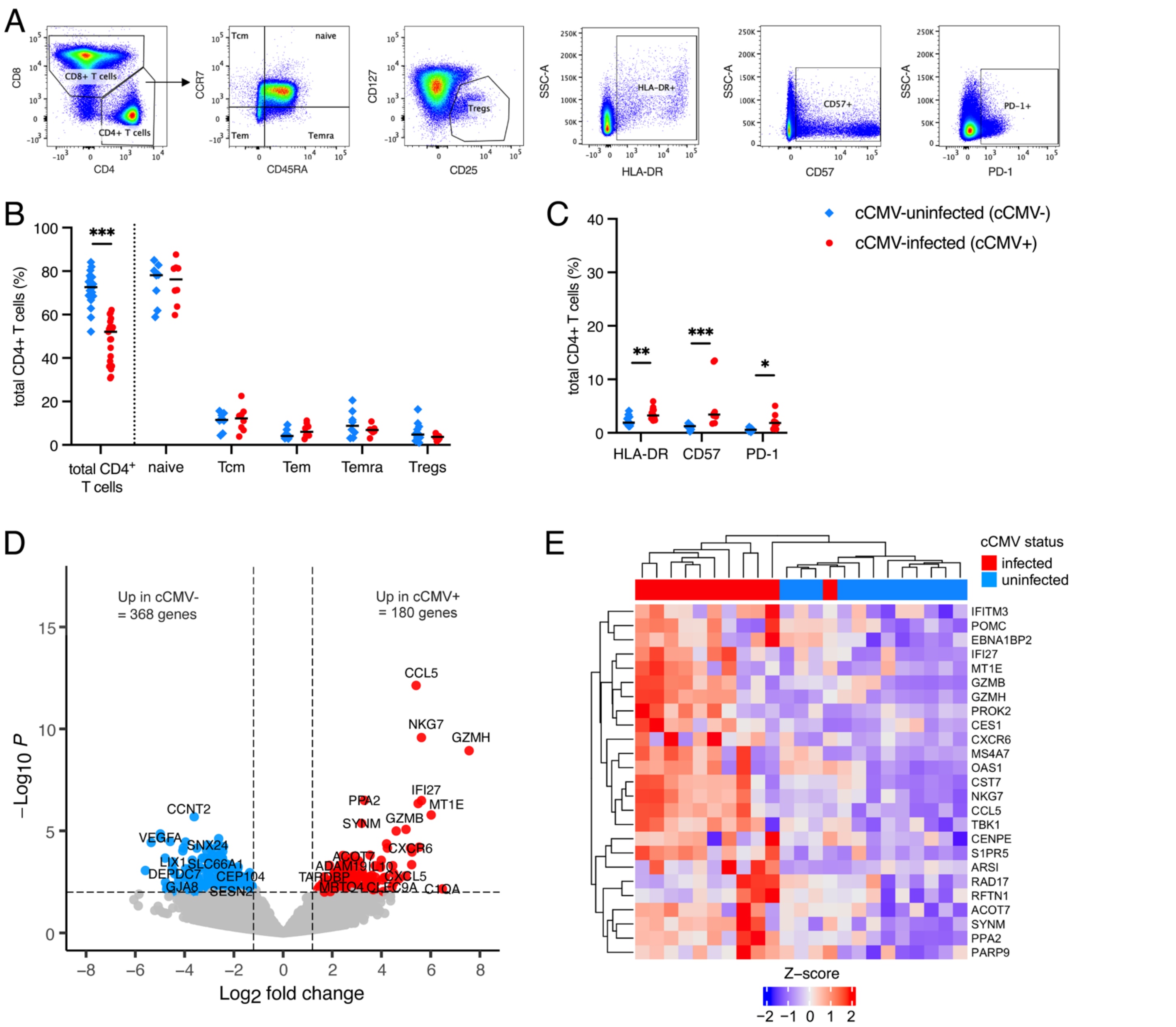


**Supplementary Figure 3. Activated CD4+ T cells increase in cord blood from cCMV-infected versus cuninfected neonates.** (A) CD4+ T cell gating strategy. (B-C) CD4+ T cell immunophenotypes were compared in umbilical cord blood from cCMV-infected (cCMV+, red circles) versus cCMV-uninfected (cCMV-, blue diamonds) neonates. (B-C) Frequency of total CD4+ T cells and CD4+ T cell subsets in cord blood from cCMV+ (n=21 total) versus cCMV- (n=20 total) neonates. (B) Frequency of total, naïve, central memory (Tcm), effector memory (Tem), terminally differentiated effector memory cells re-expressing CD45RA (Temra), and regulatory T cells (Tregs) in cord blood from cCMV+ versus cCMV- neonates. (C) Frequency of total CD4+ T cells expressing HLA-DR, CD57, and PD-1. (D) Volcano plot demonstrating differentially expressed genes in FAC-sorted total CD4+ T cells from cCMV+ (n=11) and cCMV- (n=12) neonates. Significance was set at $P$ <0.01 and log2foldchange +/-1.2. Red circles indicate genes enriched in cCMV+ CD4+ T cells, blue circles indicate genes enriched in cCMV- CD4+ T cells, and grey circles indicate genes whose expression did not differ significantly between groups. (E) Heatmap of top 25 enriched genes (FDR $P$ <0.1, log2foldchange >1.2) in CD4+ T cells from cCMV+ (n=11) versus cCMV- (n=12) neonates. Z-score shows gene expression based on rlog-transformed data. FDR-corrected $P$ values reported for Mann-Whitney U test. *$P$ < 0.05, **$P$ < 0.01, ***$P$ < 0.001.

**Supplementary Table 2. Gene set enrichment analysis (GSEA) comparing transcriptome of sorted cord blood CD8+ T cells from cCMV-infected versus uninfected neonates[a]**

| Direction | adjusted *P* value | Genes | Gene set enrichment analysis (GSEA) pathways |
|---|---|---|---|
| Down | 4.90E-03 | 22 | Regulation of axon extension involved in axon guidance |
| Up | 1.30E-03 | 49 | Chemokine-mediated signaling pathway |
| | 1.30E-03 | 57 | Response to chemokine |
| | 1.30E-03 | 57 | Cellular response to chemokine |
| | 1.30E-03 | 131 | Cellular response to interferon-gamma |
| | 1.30E-03 | 57 | Chromosome condensation |
| | 1.30E-03 | 33 | Monocyte chemotaxis |
| | 1.30E-03 | 32 | Lymphocyte chemotaxis |
| | 1.30E-03 | 115 | Nucleosome assembly |
| | 1.30E-03 | 175 | DNA packaging |
| | 1.30E-03 | 31 | DNA replication-dependent nucleosome assembly |
| | 1.30E-03 | 31 | DNA replication-dependent nucleosome organization |
| | 1.30E-03 | 53 | Natural killer cell mediated immunity |
| | 1.30E-03 | 75 | Regulation of megakaryocyte differentiation |
| | 1.30E-03 | 34 | Regulation of natural killer cell mediated immunity |
| | 1.30E-03 | 50 | Natural killer cell mediated cytotoxicity |
| | 2.10E-03 | 37 | Nucleolar chromatin organization |
| | 1.30E-03 | 31 | Regulation of natural killer cell mediated cytotoxicity |
| | 2.10E-03 | 36 | DNA heterochromatin assembly |
| | 1.30E-03 | 42 | Nucleolus organization |
| | 1.30E-03 | 88 | Leukocyte mediated cytotoxicity |
| | 1.30E-03 | 149 | Response to interferon-gamma |
| | 2.90E-03 | 25 | Positive regulation of natural killer cell mediated immunity |
| | 1.30E-03 | 126 | Cell killing |
| | 1.30E-03 | 154 | Nucleosome organization |
| | 1.30E-03 | 135 | Chromatin assembly |
| | 3.00E-03 | 13 | Positive regulation of lymphocyte chemotaxis |
| | 3.60E-03 | 16 | Pyroptosis |
| | 5.80E-03 | 21 | Positive regulation of natural killer cell mediated cytotoxicity |
| | 2.30E-03 | 10 | Eosinophil migration |

cCMV = congenital CMV infection

[a] includes n=13 cCMV-infected (cCMV+) and n=12 cCMV-uninfected (cCMV-) neonates

[b] pathways involved in natural killer (NK) cell responses highlighted in yellow

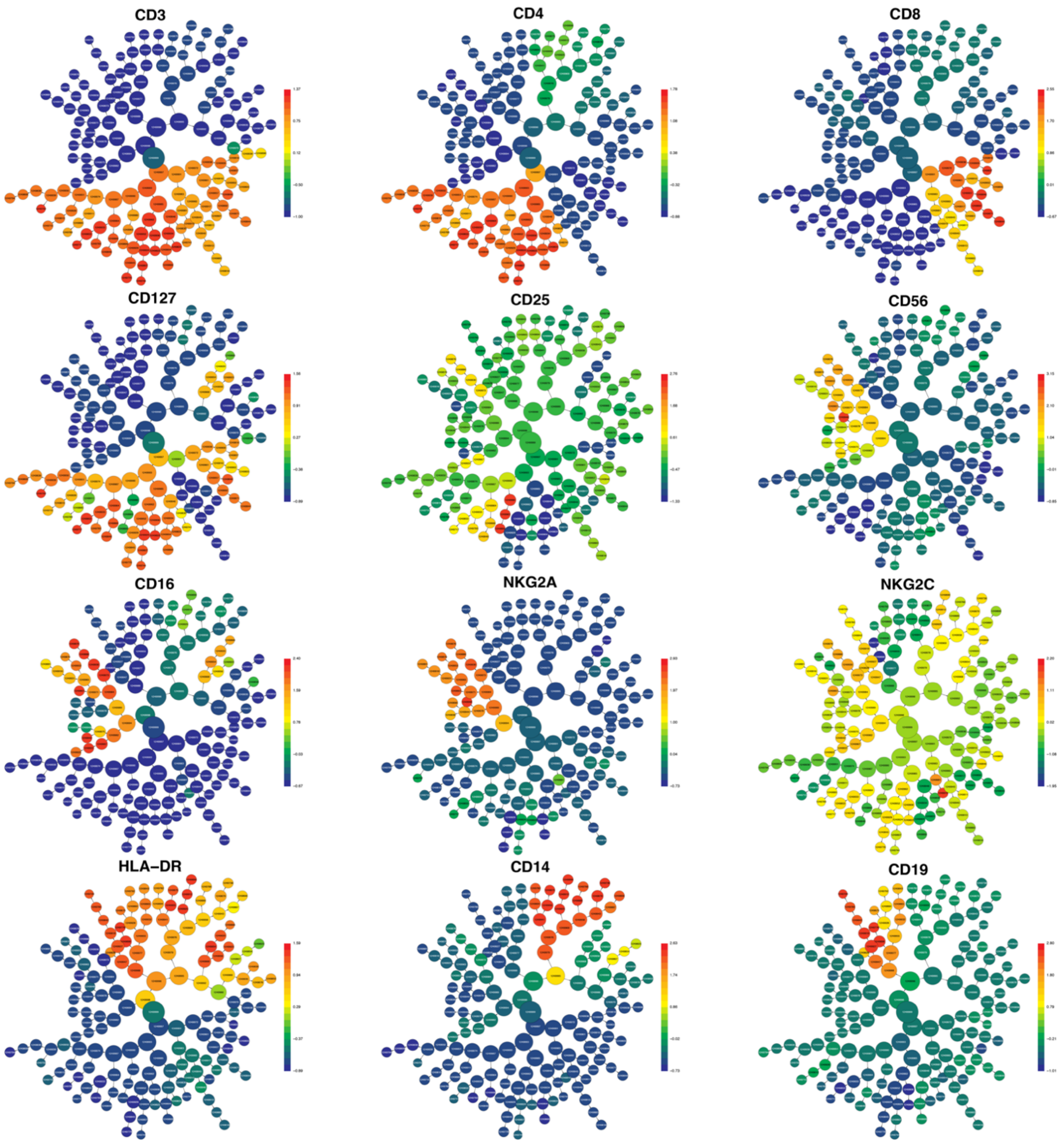


**Supplementary Figure 4. CITRUS cluster map for general lineage phenotyping panel.** Unsupervised cluster identification, characterization, and regression (CITRUS) was used to cluster immune cell populations based on CD3, CD4, CD8, CD127, CD25, CD19, CD56, CD16, NKG2A, NKG2C, HLA-DR and CD14 marker expression. CITRUS cluster map generated from 1,250,000 live, CD235a- singlet events including 50,000 events per sample (n=25 samples) with a minimum cluster size of 12,500 events (MCS = 1% of total events).

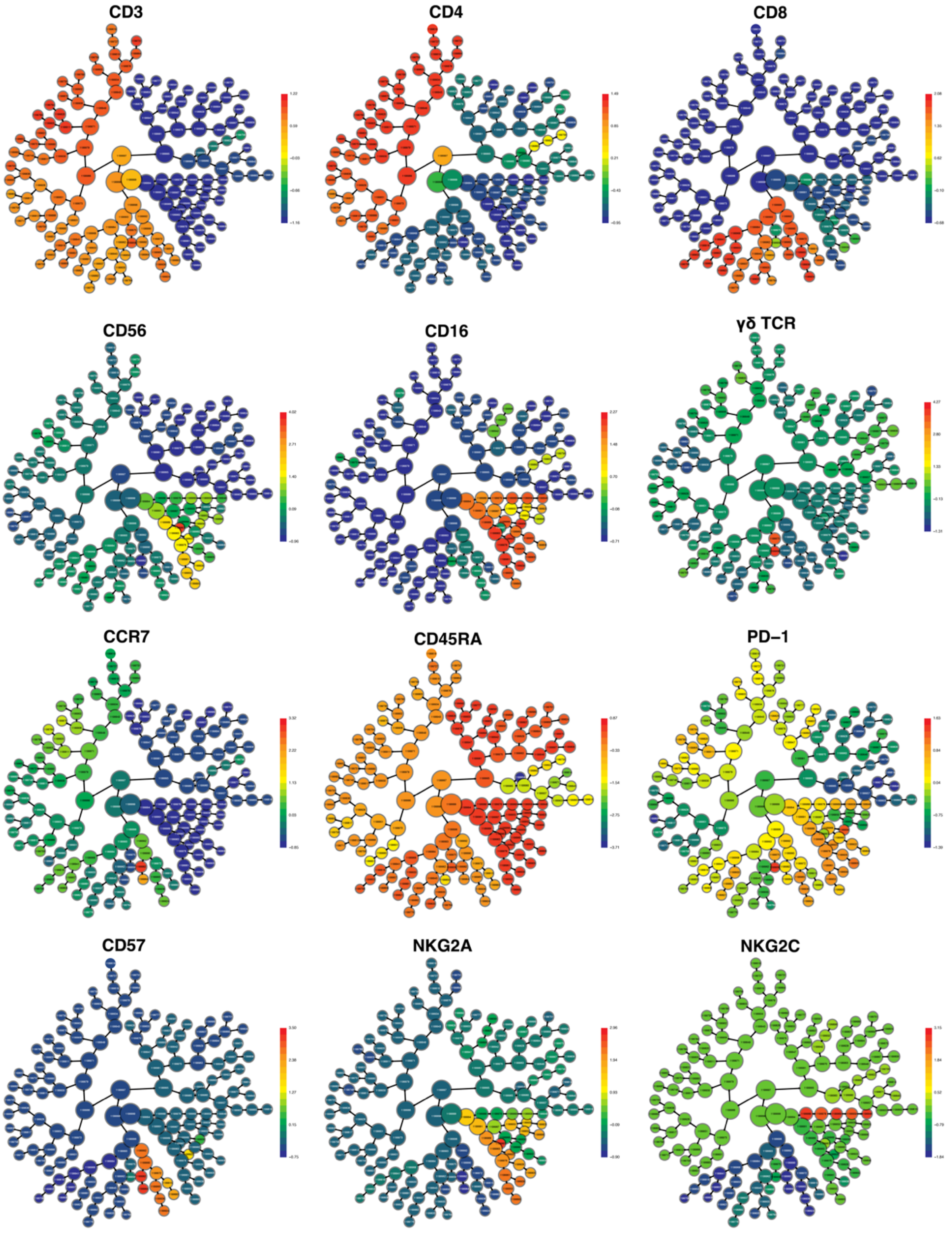


**Supplementary Figure 5. CITRUS cluster map for T and NK cell phenotyping panel.** Unsupervised cluster identification, characterization, and regression (CITRUS) was used to cluster immune cell populations based on CD3, CD4, CD8, CD56, CD16, γδ TCR, CCR7, CD45RA, PD-1, CD57, NKG2A, and NKG2C marker expression. CITRUS cluster map generated from 1,125,000 live, CD235a- singlet events including 75,000 events per sample (n=15 samples) with a minimum cluster size of 11,250 events (MCS = 1% of total events).

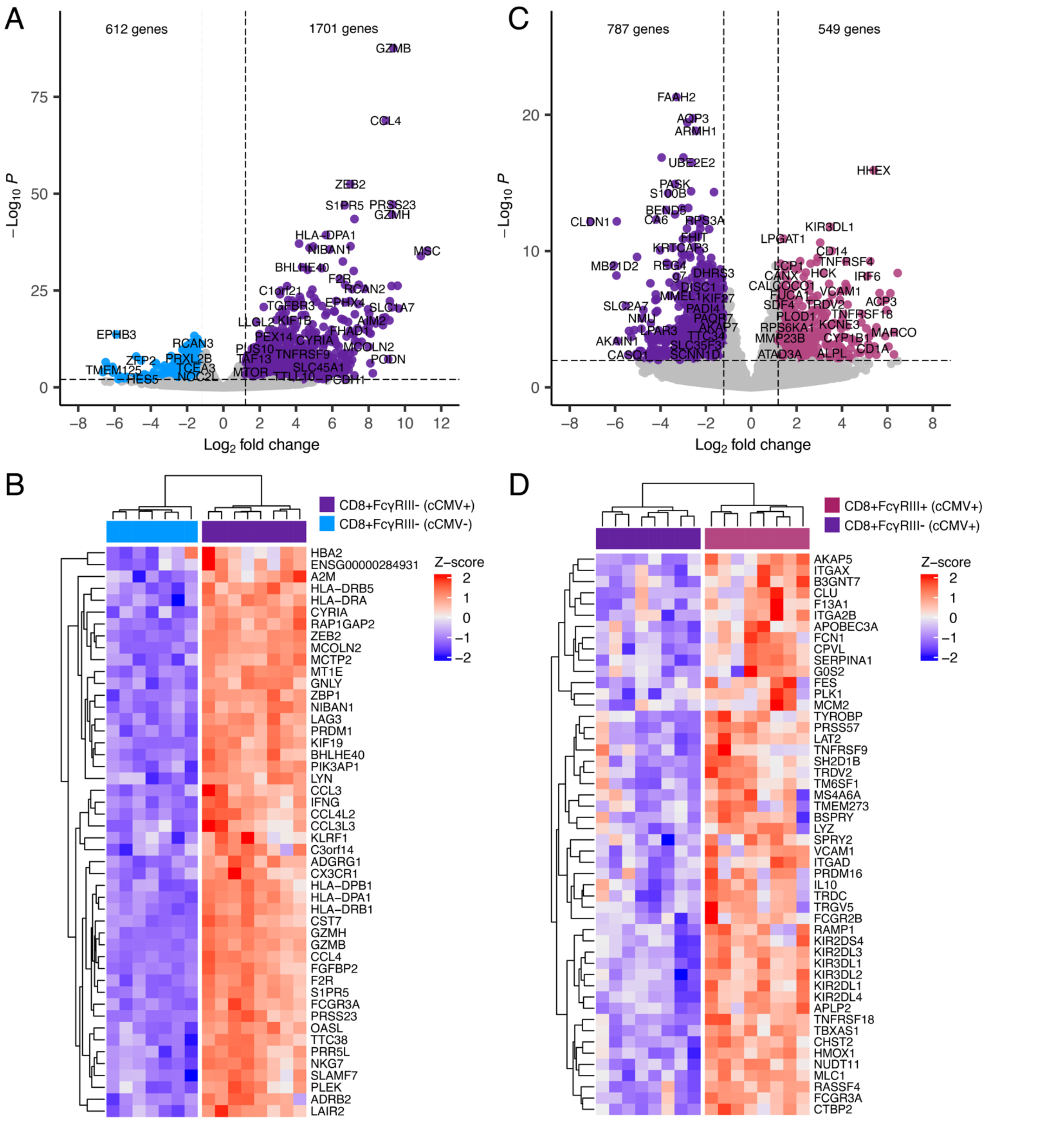


**Supplementary Figure 6. Transcriptome of FcγRIII+ and FcγRIII- CD8+ T cells in cord blood from cCMV-infected and uninfected neonates.** (A-E) Transcriptome analysis of FAC-sorted FcγRIII+ and FcγRIII- CD8+ T cells from cCMV-infected (n=8) and uninfected (n=7) neonates. (A) Volcano plot demonstrating differentially expressed genes in FcγRIII- CD8+ T cells from cCMV+ (dark purple circles) versus cCMV- (blue circles) neonates. (B) Heatmap showing top 48 differentially expressed genes (FDR *P* <0.1, log2foldchange >4.0, mean base count >500) between FcγRIII- CD8+ T cells from cCMV+ (dark purple) versus cCMV- (blue) neonates. (C) Volcano plot demonstrating differentially expressed genes in FcγRIII+ CD8+ T cells (plum circles) versus FcγRIII- CD8+ T cells (dark purple circles) from cCMV+ neonates. (D) Heatmap showing top 50 differentially expressed genes (FDR *P* <0.1, log2foldchange >2.0, mean base count >200) between FcγRIII- CD8+ T cells from cCMV+ (dark purple) versus cCMV- (blue) neonates. Significance was set at *P* <0.01 and log2foldchange +/-1.2 for volcano plots and grey circles indicate genes whose expression did not differ significantly between groups. Z-score shows gene expression based on rlog-transformed data.
